# No evidence for a causal contribution of bioavailable testosterone to ADHD in sex-combined and sex-specific two-sample Mendelian randomization studies

**DOI:** 10.1101/2023.09.09.23295037

**Authors:** Lars Dinkelbach, Triinu Peters, Corinna Grasemann, Johannes Hebebrand, Anke Hinney, Raphael Hirtz

## Abstract

The higher prevalence of attention-deficit/hyperactivity disorder (ADHD) in males raises the question of whether testosterone is implicated in ADHD risk. However, cross-sectional studies did not identify an association between ADHD and testosterone levels. Mendelian randomization (MR) studies can overcome limitations inherent to association studies, especially of reverse causation and residual confounding. In the current study, sex-combined and sex-specific two-sample MR analyses were conducted to address whether testosterone has a causal influence on ADHD risk. Sex-combined as well as sex-specific target-genetic variants for bioavailable testosterone were derived from a large genome-wide association study (GWAS) on up to 382,988 adult white European UK Biobank study participants. In our sex-specific analyses for ADHD, including data from 14,154 males and 4,945 females (17,948 and 16,246 controls respectively), no association between bioavailable testosterone and ADHD risk were found, neither in males (inverse-variance weighted (IVW): beta=0.09, 95%-CI [-0.10, 0.27]) nor in females (IVW: beta=-0.01, 95%-CI [-0.20, 0.19]). However, in the sex-combined analysis, including 38,691 cases and 186,843 controls, genetically predicted bioavailable testosterone was associated with ADHD risk (IVW: beta=0.24, 95%-CI [0.09, 0.39). The inclusion of birth weight and/or SHBG as additional variables in multivariable MR analyses did not alter this result. However, when correcting for potential BMI-driven pleiotropy by a multivariable MR study, all effect estimates for testosterone showed non-significant results. Taken together, no robust evidence for a causal effect of bioavailable testosterone on the risk for ADHD was found.

## Background

With a point prevalence of 2.2% to 7.2% attention-deficit/hyperactivity disorder (ADHD) is one of the most common psychiatric disorders in children and adolescents [29]. Current prevalence studies show a similar global proportion of 6.8% of people with symptoms persisting or originating in adulthood [33]. ADHD not only implies impaired school and work performance but is accompanied by adverse effects on health and behavior throughout the lifespan [10], accumulating in substantial disease-related costs for individuals, their families, and society [7].

Neurobiologically, disruptions in mesocortical and mesolimbic dopaminergic pathways are seen as the neurophysiologic substrate of ADHD [23], and there is conclusive evidence, especially from animal models, that the androgen testosterone modulates these pathways [18, 26]. Moreover, a potential role of testosterone in the etiology of ADHD is also suggested by the observations that ADHD is 2-3 times more prevalent in males than in females [29] and that during puberty, the symptomology of ADHD shows a distinctive change with lower impulsivity and hyperactivity but higher disease burden in adolescents and young adults [8]. Evidence for a potential role of testosterone in ADHD risk also stems from studies assessing the ratio of the second-to-fourth digit (2D:4D) length, a suggested biomarker of prenatal testosterone exposure. A recent meta-analysis of 1,405 ADHD patients found that a low 2D:4D ratio, suggestive of high prenatal testosterone exposure, was related to a higher ADHD prevalence with consistent findings by the included studies [1].

When directly measuring testosterone, one must consider that testosterone in the blood is largely bound to proteins, most importantly sex hormone-binding globulin (SHBG). Thus, only approximately 2-3% of the total testosterone is biologically active [30]. However, this bioavailable fraction is impractical to measure and thus usually approximated using equations taking total testosterone, SHBG, and levels of albumin into account [30]. In a cross-sectional study of 148 primarily prepubertal children with ADHD, levels of bioavailable testosterone were neither correlated to symptom severity nor were there significant differences between patients with ADHD and controls [36]. Studies in patients with endocrine disorders associated with increased levels of androgens provided mixed findings: While levels of testosterone were not associated with risk of ADHD in patients with congenital adrenal hyperplasia (CAH) [12], women with polycystic ovary syndrome (PCOS) scored higher on measures of hyperactivity and impulsivity [14].

Mendelian randomization (MR) studies take advantage of the random allocation of genetic variants during conception. In brief, if genetic variants (i.e., single-nucleotide polymorphisms (SNPs)) are associated with an exposure (e.g., testosterone), these genetic variants can be used to predict the expression of the exposure. An association of this genetically predicted expression of the exposure with an outcome of interest (e.g., ADHD) can provide evidence for a causal relationship between the exposure and the outcome [13]. As genetic variants are randomly allocated at birth, MR studies overcome the limitations of cross-sectional studies by avoiding reverse causation and minimizing residual confounding [13] and thus provide a reasonable approach to study the effect of genetically predicted exposure to testosterone on ADHD. In two-sample MR studies, the effect estimates are derived from two independent GWAS, allowing for even higher-powered studies.

In cross-sectional research, levels of testosterone demonstrate age-related changes, with a sharp increase during puberty and a decline later in life [11]. However, between the ages of 20 and 70, levels of testosterone tend to be comparatively stable [11]. In a sub-sample of 7,097 male and 5,285 female participants of the UK Biobank, testosterone was reassessed after approximately five years. The results showed strong correlations (r=0.68 in men and r=0.71 in women), supporting the idea of considerably stable individual baseline levels of testosterone over time [19]. Therefore, MR studies were utilized to study the effect of testosterone on various entities, including the occurrence of cancer, BMI, character traits, or fluid intelligence [19, 28].

In a previous two-sample MR study based on a GWAS of 17,666 children (aged<13 years) with ADHD, no causal effect of total testosterone, free testosterone, and SHBG on ADHD symptom severity was found [19]. However, a two-sample MR study based on a case-control study on 94,478 adult males (390 cases with ADHD) and 122,986 women (418 cases with ADHD) of the FinnGen cohort found evidence for a directional effect of total testosterone in males and SHBG in both sexes for ADHD risk [19].

Given the above outlined somewhat conflicting results, the present MR study was undertaken to assess whether bioavailable testosterone causally contributes to ADHD. The present study has greater statistical power than previous studies, as it is based on a large GWAS on 382,988 adult participants on bioavailable testosterone [28] and a recently published case-control GWAS on 38,691 children and adults with ADHD and 186,843 controls [5]. Considering the hardly overlapping genetic underpinnings of testosterone in males and females [19, 28] and the sex-specific ADHD symptomology with higher levels of hyperactivity in males [20], extensive sex-combined and sex-specific analyses were conducted.

## Methods

### Study design and assumptions

The validity of genetic variants to serve as an instrumental variable (IV) in MR studies relies on three assumptions i.) the genetic variants are related to the exposure of interest (relevance criterion), ii.) the genetic variants are not related to confounding pathways influencing the outcome (exchangeability), and iii.) the genetic variants are only related to the outcome via the exposure and not directly related to the outcome (exclusion restriction). To ensure that the first assumption was met, the genetic variants chosen in our study are based on a large-scale GWAS, with genome-wide significance at a commonly accepted threshold of p<5×10^-8^ [28]. Moreover, F-statistics were calculated to assess the strength of each SNP and for the IV. By convention, a weak instrument is defined as an instrument with an F-statistic<10. The second and third assumptions cannot be tested directly. Horizontal pleiotropy and IV heterogeneity lead to violation of assumptions for MR and biased effect estimates. To assess the degree of heterogeneity, Cochran’s Q statistic was calculated. Significant heterogeneity was addressed by a) calculating a distinctive set of methods robust against heterogeneity, b) assessing several types of pleiotropy using suitable methods, and c) by including phenotypes that are a potential source for pleiotropy in a multivariable MR (MVMR) analysis. To identify relevant phenotypes for the MVMR analysis, the PhenoScanner database [16, 34] was queried regarding all SNPs used to define the bioavailable testosterone IV (threshold: p<5×10^-8^). Among the identified phenotypes, (1) those confirmed by a comprehensive literature review, (2) those serving as overarching phenotypes that encompass additional candidate phenotypes (i.e., they represent neither upstream nor downstream mechanisms), and (3) those with available GWAS data were included for further MVMR analyses. As a result, the body mass index (BMI) [25], SHBG [19, 28], and birth weight [37] qualified as confounders.

### Statistical methods

First, effect estimates were calculated via the inverse variance weighted (IVW) method, which calculates an effect estimate of the exposure on the outcome using a simple regression model, whereby the effect of the genetic variant on the outcome is weighted by the effect of each variant on the exposure variable. Several robust methods have been developed to consider and correct for violations of the MR assumptions. According to recent recommendations, the following methods were calculated: MR-Egger, simple and weighted median, and mode-based effect estimates, the penalized weighted median method, MR-PRESSO, MR-Lasso, the contamination mixture method, and the MR-RAPS method. Additional information on the procedures and software packages used in these methods and their advantages and disadvantages can be found in the Supplementary Methods 1 and 2. Scatter plots illustrating the effect of each SNP on the exposure and outcome, funnel plots that visualize the relationship between the effect estimate of each genetic instrument and the inverse of the standard error as a measure of the instruments’ precision, and forest plots of the effect estimate of each SNP were created to screen for violations of the MR assumptions visually. Leave-one-out analyses were conducted to assess whether effects were driven by single variants. MR analyses in the present study were performed and are reported following the STROBE-MR guidelines [32].

### Study populations

#### Testosterone GWAS and testosterone measurement

The instrumental variable for bioavailable testosterone for sex-combined analyses was derived from a large GWAS on samples of 382,988 white European UK Biobank study participants aged 40 to 69 years [28]. For sex-specific analyses, instrumental variables were based on GWAS on samples of 178,782 male and 188,507 female study participants. Genetic instruments were defined based on independent variants with a genome-wide significance (p<5×10^-8^) for a given trait and sex. In the GWAS regression models, age, the genotyping chip, and ten principal components for ancestry were included as covariates (among specific covariates for individual traits). In the sex-combined and female-specific GWAS, menopause as well as the time of sample collection were included as covariates. SNP-based heritability (h²_SNP_) of bioavailable testosterone was 14% in women and 12% in men. Further details regarding the measurement of bioavailable testosterone can be found in the Supplementary Methods 3.

#### ADHD – sex-combined GWAS

For sex-combined outcome associations, summary statistics of a GWAS-meta-analysis on 38,691 participants (33.4% females) with ADHD and 186,843 controls (53.0% females) were used, which utilized data from the Danish iPSYCH cohort, the Icelandic deCODE cohort and previously published data from ten European ADHD study groups of the Psychiatric Genomics Consortium (PGC) [5]. For participants of the iPSYCH and PGC cohorts, a diagnosis of ADHD was made by psychiatrists according to ICD-10 criteria (F90.0, F90.1, F98.8 diagnoses for the iPSYCH cohort and F90.0 only for the PGC cohort). For participants of the deCODE cohort, participants with ADHD were either identified by a diagnosis of ADHD (F90.0, F90.1, and F98.8 (ICD-10)) or by the prescription of ADHD medication (mostly methylphenidate). In addition, probabilities (i.e., dosages) resulting from the imputation of genotypes, regression models used an additive logistic regression with the first ten principal components for the iPSYCH cohort and sex, year of birth, and county of origin as covariates for the deCODE cohort. SNP-based heritability (h²_SNP_) was 14%.

#### ADHD - sex-specific GWAS

For sex-specific outcomes, summary data of a case-control GWAS on 14,154 males and 4,945 females of European ancestry with ADHD (with 17,948 male and 16,246 female controls) were used [21]. The population of this GWAS utilized earlier waves of the iPSYCH and PGC cohorts as described above and thus largely overlaps with the GWAS used in the sex-combined analyses but provides sex-specific summary statistics. No details on the covariates included in the GWAS were given [21]. SNP-based heritability (h²_SNP_) was 12.3% for females and 24.7% for males. Table 1 gives an overview of the study populations utilized in our univariable MR analyses.

**Table 1.** Exposure and outcome GWAS used in sex-combined and sex-specific univariable MR studies.

| sex | Exposure GWAS (bioavailable testosterone) |  |  |  | Outcome GWAS (ADHD) |  |  | 80% Power<br>OR per SD<br>exposure |  |
| --- | --- | --- | --- | --- | --- | --- | --- | --- | --- |
|  | study | N | SNPs | PVE | study | cases | controls |  |  |
| sex-combined | Ruth et al., 2020 | 382,988 | 147 | 3.5% | Demontis et al., 2023 | 38,691 | 186,843 | $\leq 0.918$ | $\geq 1.085$ |
| male | Ruth et al., 2020 | 178,782 | 125 | 7.9% | Martin et al., 2018 | 14,154 | 17,948 | $\leq 0.894$ | $\geq 1.117$ |
| female | Ruth et al., 2020 | 188,507 | 180 | 6.9% | Martin et al., 2018 | 4,945 | 16,246 | $\leq 0.835$ | $\geq 1.178$ |

### Multivariable MR analyses

Multivariable MR (MVMR) is an extension of the univariable MR approach to simultaneously estimate the effect of several exposures (i.e., bioavailable testosterone as well as the abovementioned confounders BMI, SHBG, and birth weight) on the outcome of interest (ADHD) [2]. Only index-SNPs with available data in all the included GWASs were considered. For MVMR analyses, multivariable extensions of the abovementioned IVW, MR-EGGER, the median method, the MR-Lasso method, and MR-PRESSO were calculated. Details on the study populations and the GWAS used in the MVMR studies are given in the Supplementary Methods 4.

### Instrumental variables

For SNPs not available in the outcome GWASs, proxy SNPs were searched for the conduction of univariable MR studies. For sex-specific analyses, proxy-SNPs as suggested by Ruth et al. [28] with a r²>0.5 (based on HapMap2) were used. For sex-combined analyses, 13 proxy SNPs with a r²>0.5 were identified using LDlink [24]. Due to a limited number of identified proxies, the search was extended with the same r² threshold (>0.5) by directly querying the Ensembl REST API [4], which resulted in 10 additional proxy-SNPs. In case of the availability of multiple proxy SNPs, the SNPs were chosen according to (first) r^2^ and (second) their distance to the queried SNP. A PhenoScanner search for missing SNPs without an identified proxy did not indicate that a major phenotype association was missed (see Supplemental Table S3). Analyses of primary endpoints were repeated with proxies that were selected with a threshold of r²>0.8 as part of the sensitivity analyses.

#### Ethics

In all the GWAS that were utilized in this study, all participants (or their legal guardians) provided written informed consent and approval from the local ethics committees were obtained prior to data collection.

## Results

### Sex-combined analyses

Eighty-seven of 147 index SNPs for the exposure “sex-combined bioavailable testosterone in both sexes” [28] were available in the sex-combined GWAS on ADHD [5]. Twenty-three of the missing SNPs could be replaced by proxy SNPs (see Supplemental Table S1 for details). MR-PRESSO identified three pleiotropic SNPs (rs1811450, rs28929474, and rs4431046, see Supplementary Results 1), which were excluded from further analyses, resulting in a total of 107 SNPs defining the IV. No weak instruments were included in the analyses (median F-statistic per SNP 47.34, range 19.74 to 756.95). The F-statistic of the instrumental variable was 8180.99. The IVW method showed an association between bioavailable testosterone and ADHD (b=0.24, p=0.002). However, the Q-statistic revealed significant heterogeneity (IVW: Q-statistic: df=106, 187.58, p=1.79×10^-6^; MR-Egger: df=105, Q-statistic: 185.64, p=2.06×10^-6^). Robust methods addressing heterogeneity showed a mixed picture. The MR-Lasso method revealed a significant effect estimate (b=0.24, p=1.78×10^-4^), just as the simple median-based estimator was significant (b=0.25, p=0.010), and MR-RAPS (b=0.24, p=0.003). All other median- and mode-based methods provided positive effect estimates but were non-significant, consistent with the results from MR-Egger analysis, even with bootstrapping (Figure 1). The funnel plot, scatter plot, leave-one-out analysis, as well as the corresponding forest plot did not show any violation of MR assumptions or effects that were driven by single SNPs (Supplemental Figures S1-S3).

**Figure 1.**
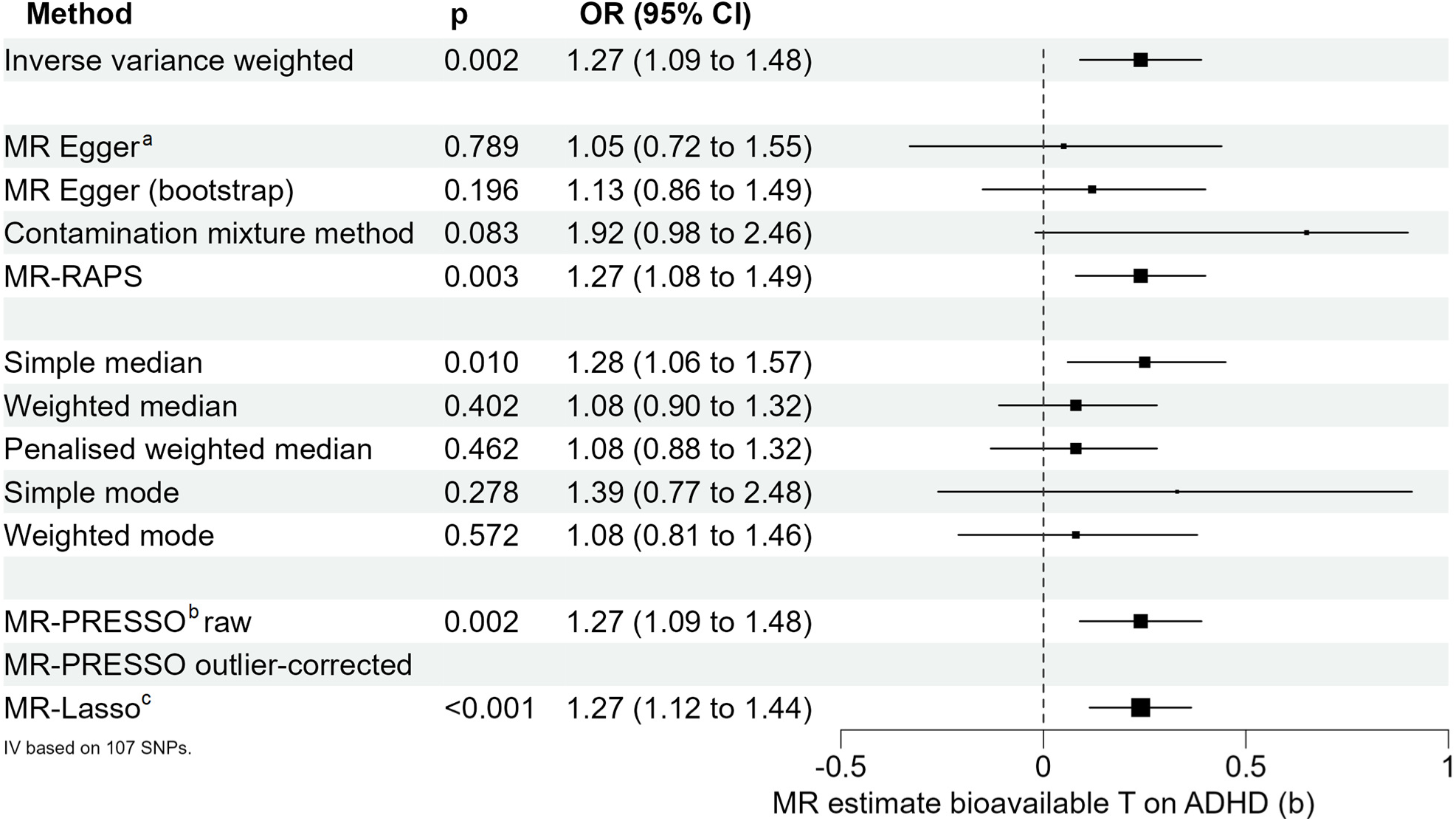
Univariable MR analyses on the effect of bioavailable testosterone on ADHD integrating both sexes. ^a^ Eggers-intercept did not show evidence for significant directional pleiotropy (intercept=0.0038, se=0.0036, p=0.298). ^b^ The MR-PRESSO global test for pleiotropy was significant (191.69, p<3.3×10^-4^). No additional outliers were identified after the exclusion of the three outliers identified by the MR-PRESSO method. Thus, no outlier-corrected MR estimate was calculated. ^c^ The MR-Lasso method identified 90 valid instruments (tuning parameter=0.183).

#### Sensitivity analyses

To test whether the r^2^ threshold for proxy SNPs, the harmonization procedure, or the exclusion of SNPs by MR-PRESSO biased the results of our study, two additional analyses were conducted, which, overall, replicated the results of the main analysis (Supplemental Figures S10 to S11).

#### Phenotype associations

The PhenoScanner search revealed several phenotype associations of the genetic variants used in the sex-combined MR analysis (see Supplemental Table S2 for details). For 59 of the 110 queried SNPs, the PhenoScanner search provided hits. Direct upstream/downstream mechanisms of bioavailable testosterone affecting other androgens (e.g., dihydrotestosterone or dehydroepiandrosterone-sulfate, n=5 SNPs), height (n=27 SNPs), bone density (n=4 SNPs), and pubertal timing or menstrual cycle (n=9 SNPs) were interpreted as unlikely pleiotropic influence on the testosterone∼ADHD association. Another large group of SNPs was related to weight and/or BMI (n=17 SNPs), with a reasonable effect on ADHD (see Supplemental Methods 4 for details), metabolic syndrome-related alterations, e.g., lipoprotein levels or insulin resistance (n=16 SNPs), or cardiovascular diseases (n=10 SNPs), all having strong associations with BMI. N=21 SNPs were related to blood count alterations, including all three cell lines (erythrocytes, leukocytes, and platelets), presumably unrelated to ADHD.

#### MVMR analyses

Given significant heterogeneity (Q-Test results) and evidence for pleiotropy (MR-PRESSO) in univariable MR analyses, multivariable MR analyses were conducted, including BMI, birth weight, and SHBG, as suggested by the PhenoScanner search and the current state of literature (for details on the rationale behind exposure variable selection see Supplementary Methods 1). As data were not available on an individual-level, phenotype correlations between exposure variables could not be considered. To consider the relationship between BMI and birth weight, two separate MVMR analyses were conducted, one with BMI and one with birth weight, including the remaining exposures (bioavailable testosterone and BMI-adjusted SHBG) in each of these MVMR studies. For MVMR, the same instrumental variable consisting of 110 genetic variants as for the univariable MR analysis was used (including proxies, see Supplemental Tables S4 and S5).

In the first MVMR model, MR-PRESSO identified two pleiotropic SNPs (rs1811450, rs7496293, see Supplementary Results 6), which were excluded from further analyses, resulting in 108 SNPs defining the IV. F-statistics were 24.92 for bioavailable testosterone, 54.25 for BMI-adjusted SHBG, and 6.68 for BMI. The Q-test yielded a significant result (df=104, Q=187.59, p=9.67×10^-7^). Including SHBG and BMI, all effect estimates for bioavailable testosterone yielded non-significant results. For BMI, the IVW method, MR-Egger, and MR-Lasso yielded significant findings. For SHBG, all effect estimates were non-significant (Figure 2A).

**Figure 2.**
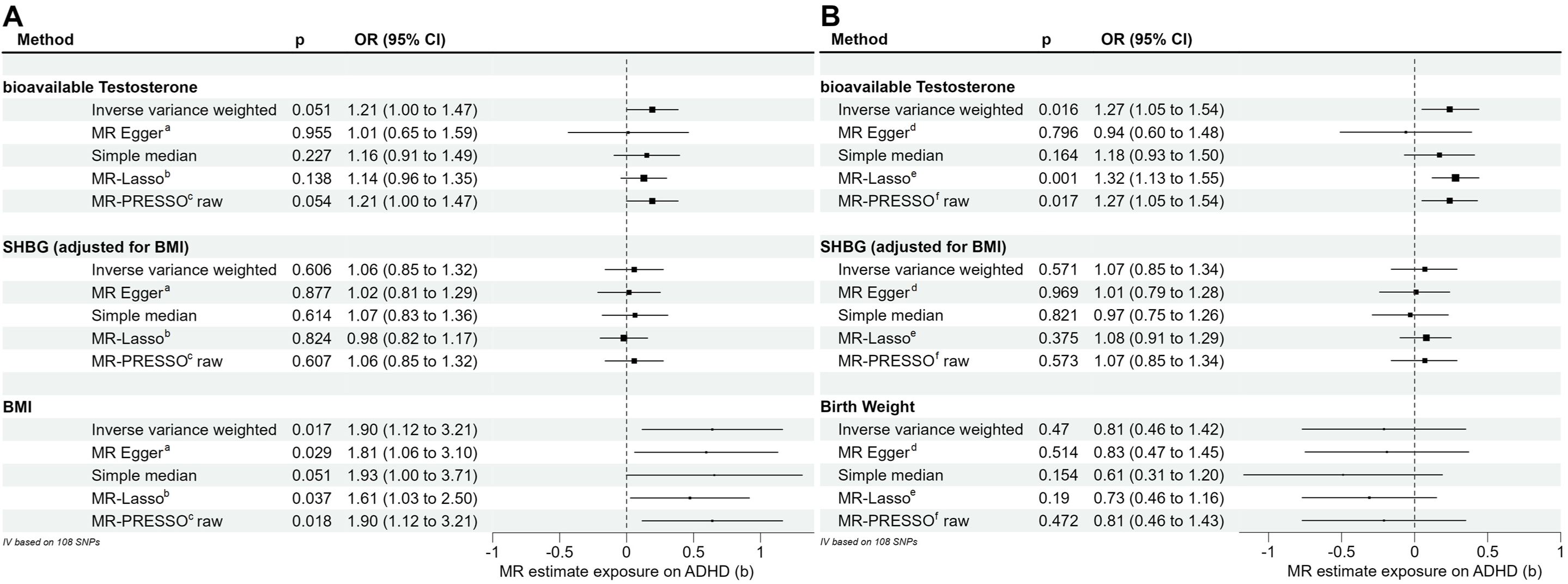
Results of the multivariable sex-combined MR analyses on ADHD. Figure 2A depicts the results of the MVMR analysis with bioavailable testosterone, BMI-adjusted SHBG, and BMI. Figure 2B depicts MVMR analysis with bioavailable testosterone, BMI-adjusted SHBG and birth weight. ^a^ Eggers-intercept did not show evidence for significant directional pleiotropy (intercept=0.003, se=0.004, p=0.386). ^b^ The MR-Lasso method identified 85 valid instruments (tuning parameter=0.165). ^c, f^ After the exclusion of the two outliers identified by the MR-PRESSO method, no additional outliers were identified. Thus, no outlier-corrected MR estimate was calculated. ^d^ Eggers-intercept=0.006, se=0.004, p=0.149. ^e^ The MR-Lasso method identified 90 valid instruments (tuning parameter=0.177).

For the second MVMR model, including bioavailable testosterone, BMI-adjusted SHBG, and birth weight as exposure variables, MR-PRESSO identified two pleiotropic SNPs (rs1811450, rs4431046, see Supplementary Results 6 for details) that were excluded from further analysis, resulting in 108 SNPs defining the IV. The F-statistics were 18.99, 17.12, and 3.05, respectively. The Q-statistic was significant (df=105, Q=195.4, p=1.49×10^-7^). The IVW and MR-Lasso methods revealed significant findings for bioavailable testosterone, while the MR-Egger and median method were not significant. All effect estimates for SHBG and birth weight were not significant (Figure 2B).

As a sensitivity analysis, both MVMR models were recalculated without the exclusion of pleiotropic SNPs based on MR-PRESSO, resulting in similar effect estimates as the primary analyses (Supplementary Results 6 and Supplemental Figure S14).

### Sex-specific analyses

#### Males

Seventy-nine of 125 index SNPs for the exposure “bioavailable testosterone in males” [28] were available in the GWAS on ADHD by Martin, et al. [21]. Of the missing SNPs, 10 could be replaced by proxy SNPs (Table S6). The F-statistic of the instrumental variable was 2759.21. No weak instruments were included in the analysis (range 21.8 to 413.4). The IVW did not provide evidence for an effect of bioavailable testosterone on ADHD in males (b=0.09, p=0.354). The Q-statistic revealed some degree of heterogeneity (IVW: df=88, Q=149.4, p=4.9×10^-5^; MR-Egger: df=87, Q=148.2, p=4.7×10^-5^). Only the bootstrapped MR-Egger method provided a significant result (b=0.29, p=0.042); however, without bootstrapping, the effect estimate of MR-Egger was not significant. All further robust methods provided non-significant results (Figure 3A, Supplemental Figures S4-S6).

**Figure 3.**
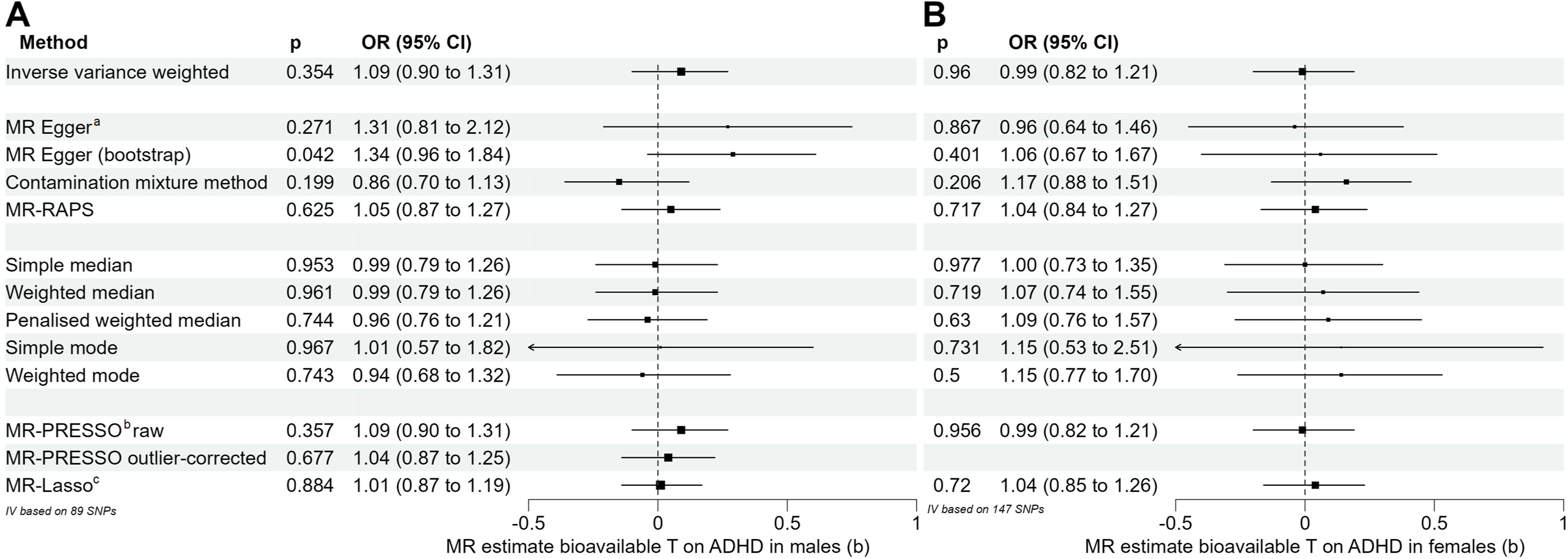
Results of the sex-specific MR analysis on the effect of bioavailable testosterone on ADHD. Figures 3A and 3B illustrate the effect estimates for the analysis in males and females, respectively. ^a^ Eggers intercept did not show any evidence for significant directional pleiotropy, neither in males (intercept=-0.006, se=0.0078, p=0.417) nor in females (intercept=0.0008, se=0.0052, p=0.872). ^b^ The MR-PRESSO global test for pleiotropy was significant in males (153.159, p=5×10^-4^) but not significant in females (150.876, p=0.405). ^c^ The MR-Lasso method led to the identification of 75 valid instruments in males (tuning parameter=0.209) and 146 valid instruments in females (tuning parameter=0.227).

#### Females

A total of 129 of 180 index SNPs for the exposure “bioavailable testosterone in females” [28] were available in the GWAS on ADHD by Martin, et al. [21]. Of the missing SNPs, 18 were replaced by proxy SNPs (Supplemental Table S7). The F-statistic of the instrumental variable was 1590.44. No weak instruments were included in the analysis (median F-statistic per SNP 36.0, range 18.8 to 1002.8). The IVW did not provide evidence for an effect of bioavailable testosterone on the risk for ADHD in females. All further methods provided non-significant results (Figure 3B, Supplemental Figures S7-S9). Q-statistic-based tests for heterogeneity were non-significant (IVW: df=146, Q=148.91, p=0.417, MR-Egger: df=145, Q=148.89, p=0.395).

#### Sensitivity analyses

The effect-allele frequency was not provided in the summary statistics for sex-specific GWAS on ADHD. Therefore, the effect-allele frequency could not be utilized for the harmonization of palindromic SNPs in sex-specific analyses. We recalculated the sex-specific MR analyses with instrumental variables excluding palindromic SNPs, which overall replicated our findings (Supplemental Results 5).

## Discussion

Several studies indicate a relationship between testosterone and AHDH. However, conclusions from these studies are limited by their cross-sectional design or other methodological constraints. For this reason, we conducted multiple MR studies and performed sex-combined MR analyses with MR studies addressing sex-specific effects, while additionally considering potential sources of pleiotropy. Overall, no robust effect of bioavailable testosterone on ADHD could be found.

The sex-combined MR study relied on a meta-analysis including three study groups with a total of 38,691 participants with ADHD and 186,843 controls. Considering significant evidence of heterogeneity in this analysis, several robust methods accounting for different sources of pleiotropy turned out to be non-significant. An analysis of the phenotypic associations of the genetic variants utilized in the MR study showed that several genetic variants were either directly or indirectly associated with body weight, obesity and/or birth weight. The inclusion of BMI and birth weight (together with BMI-adjusted SHBG) in two separate MVMR analyses showed that BMI but not birth weight led to non-significant effect estimates for bioavailable testosterone on ADHD risk. Thus, altogether, there was no evidence of a causal contribution of bioavailable testosterone to the susceptibility to ADHD. This was also found by the sex-specific analyses based on data from 14,154 men and 4,945 women with ADHD, which however were less well-powered, especially the analyses in females.

Our results are in line with the findings from a cross-sectional study in 148 mainly prepubertal boys and girls and 72 controls which found neither a significant difference in bioavailable testosterone between patients and controls nor a significant correlation of bioavailable testosterone with symptom severity in ADHD patients [36]. In an epidemiological analysis using insurance data of patients with CAH, a condition that is related to increased intrauterine and often postnatal androgen levels, no evidence for a higher prevalence of ADHD was found in comparison to matched subjects without CAH [12]. In contrast, a recent meta-analysis pooling evidence from nine studies including 1,405 participants found an association between the 2D:4D length ratio and ADHD risk as well as the severity of hyperactive/inattentional symptoms [1]. However, while the 2D:4D ratio is often seen as a surrogate marker for intrauterine testosterone exposure, the observation of a missing association of testosterone levels in amniotic fluid with the 2D:4D ratio only recently failed to confirm this idea [27], a finding questioning the validity of this measure. In addition, it has been shown that the 2D:4D length ratio varies, e.g., with maternal smoking [31] or birth weight [17], which are both not per se associated with intrauterine testosterone exposure.

Concerning the postnatal androgen status, a study of 40 women with polycystic ovary syndrome (PCOS) showed a higher burden of hyperactive/impulsive symptoms in women with PCOS than in controls [14]. However, PCOS has a considerable heritable component [35] and is strongly associated with higher BMI levels, both factors were not accounted in the study.

A previous publication utilized MR to study the effect of testosterone on ADHD risk as part of a large trial on the effect of testosterone on several somatic as well as psychiatric phenotypes [19]. In sex-specific analyses, including a very limited sample of 390 males and 418 females with ADHD, a positive association between total testosterone and risk for ADHD in males was found [19]. However, total testosterone does not provide an estimate of the biologically active testosterone fraction and is therefore not recommended to be used for the assessment of clinical effects of testosterone [30].

In the multivariable MR analysis, BMI was found to exert a causal effect on ADHD risk. This finding is consistent with a bidirectional one-sample MR study in 7,446 men that provides suggestive evidence for a causal effect of BMI on testosterone, with higher BMI levels leading to lower testosterone levels [9]. Additionally, obesity in children and adolescents has repeatedly been linked to a higher ADHD risk [3]. A bidirectional MR study supports this observation and found evidence for BMI driving an increased ADHD risk rather than ADHD leading to a rise in BMI [22].

### Limitations

Our study has some limitations. First, the definition of the exposure variable, bioavailable testosterone, was based on a single-point measurement in adult participants [28]. Testosterone levels vary with age, time of sample collection, and, in women, with menopause [6, 11], and thus, these variables have been accounted for as covariates in the GWAS used in our study. However, given the peripubertal dynamics of testosterone, inferences regarding testosterone levels during childhood and adolescence from the GWAS used in this study must be drawn with caution, even though lasting effects of genetic variants associated with bioavailable testosterone levels seem plausible. Second, due to limited overlap of SNPs in the exposure and outcome GWAS, the proportion of index SNPs that could be included as instrumental variable was ∼70-80% in all the performed MR analyses, despite incorporation of available proxy SNPs. Multiple sensitivity analyses did not indicate a bias by these limitations. Third, the sex-specific analyses, especially in females, were afflicted by low power due to a sample size of 4,945 patients with ADHD. Lastly, a polymorphism of the androgen receptor gene with a known modulating effect of testosterone, a variable number of CAG and GGN repeat is not available on the GWAS chips, and thus could not be considered in the present analyses. However, this SNP has been shown to affect the relationship between testosterone levels and depression severity in adolescents with depression [15].

## Conclusions

There is no evidence for a causal contribution of bioavailable testosterone to the risk for ADHD in this MR study. BMI is a potential confounder of the testosterone-ADHD relationship and should be considered in studies addressing this relationship. Future studies should address whether testosterone within specific, sensitive windows during brain development (e.g., fetal period, childhood, or adolescence) and/or polymorphisms of the androgen receptor play a potential role in ADHD risk.

## Supporting information

Supplementary Methods and Results

## Data Availability

All data produced in the present study are available upon reasonable request to the authors.

https://static-content.springer.com/esm/art%3A10.1038%2Fs41591-020-0751-5/MediaObjects/41591_2020_751_MOESM3_ESM.xlsx

https://figshare.com/articles/dataset/adhd2022/22564390

https://figshare.com/articles/dataset/adhdSexSpecific2018/19383299

https://www.ebi.ac.uk/gwas/studies/GCST009004

http://egg-consortium.org/birth-weight-2019.html

## Abbreviations

2D:4D: second-to-fourth digit length
ADHD: attention-deficit/hyperactivity disorder
BMI: body mass index
CAH: congenital adrenal hyperplasia
GWAS: genome-wide association study
IVW: inverse-variance weighted
MAF: minor allele frequency
MR: Mendelian randomization
MVMR: multivariable Mendelian randomization
PCOS: polycystic ovary syndrome
SHBG: sex hormone-binding globulin
SNP: single-nucleotide polymorphism

## Author contributions

TP and RH conceptualized the study. TP and RH designed and supervised the statistical analyses. LD conducted the statistical analyses. LD wrote the first draft of the manuscript. TP, CG, JH, AH, and RH critically revised the manuscript.

