## Supplementary Methods and Results for "No evidence for a causal contribution of bioavailable testosterone to ADHD in sex-combined and sex-specific two-sample Mendelian randomization studies"

### Overview

### Supplementary Methods 1 – Statistical methods to calculate effect estimates

### Supplementary Methods 2 – Software

### Supplementary Methods 3 – Measurement/calculation of bioavailable testosterone

### Supplementary Methods 4 – Exposure variables and populations included in MVMR studies

### Supplementary Results 1 – MR study on bioavailable testosterone on ADHD (sexes-combined)

### Supplementary Figure S1 – Scatter Plot

### Supplementary Figure S2 – Funnel Plot

### Supplementary Figure S3 – Forest Plot

### Supplementary Results 2 – MR study on bioavailable testosterone on ADHD (males only)

### Supplementary Figure S4 – Scatter Plot

### Supplementary Figure S5 – Funnel Plot

### Supplementary Figure S6 – Forest Plot

### Supplementary Results 3 – MR study on bioavailable testosterone on ADHD (females only)

### Supplementary Figure S7 – Scatter Plot

### Supplementary Figure S8 – Funnel Plot

### Supplementary Figure S9 – Forest Plot

### Supplementary Results 4 – Sensitivity analyses (sexes-combined analyses)

### Supplementary Figure S10 – sexes-combined MR analysis with proxy selection based on r²>0.8

### Supplementary Figure S11 –sexes-combined MR analysis with exclusion of palindromic SNPs with intermediate allele frequencies.

### Supplementary Results 5 – Sensitivity analyses (sex-specific analyses)

### Supplementary Figure S12 – males-only MR analysis with exclusion of palindromic SNPs.

### Supplementary Figure S13 –females-only MR analysis with exclusion of palindromic SNPs.

### Supplementary Results 6 – Sensitivity analyses (MVMR)

### Supplementary Figure S14 – MVMR analysis without exclusion of pleiotropic SNPs.

### Supplementary References

### Supplementary Methods

This section provides details on the methodology and its rationale used in the current study.

**Methods 1 - Statistical methods to calculate effect estimates**

As primary outcome, effect estimates were calculated via the Inverse Variance Weighted (IVW) method which calculates an effect estimate of the exposure on the outcome using a simple regression model, whereby the effect of the genetic variant on the outcome is weighted by the effect of each variant on the exposure variable. This approach relies on three assumptions: i.) all genetic variants are valid instrumental variants, i.e. fulfilling the MR-assumptions, ii.) pleitropic effects are independent of the instrument strength, i.e. the beta weight of the instrument on the exposure (so called InSIDE assumption), iii.) the mean pleiotropic effect of all instrumental variables is zero. Eggers intercept is calculated to assess the amount of directional pleiotropy. To consider and correct for violations of these assumptions, several so-called robust methods have been developed. These methods differ in their robustness and ability to correct for the source and degree to which these assumptions are violated. As it is recommended to calculate several methods to benefit from their specific advantages, the following set of robust methods were calculated: 1.) The MR-Egger method introduces an intercept to the regression model, allowing for directional pleiotropy (pleiotropic effect ≠ 0), however relies on the InSIDE assumption [2]. 2-5.) Median- and mode-based methods rely on the median (or mode) of ratio estimates and thus are more robust to (potentially pleiotropic) outliers and therefore only at least 50% (for median based methods) or the majority (for mode-based methods) of instrumental variants must be valid instruments. However, median- or mode-based methods are usually less effective (smaller power). Either simple (with all genetic variants receive equal weights in the analysis) or weighted (with each variant being weighted similarly as the weights used in the IVW method) median- and mode-based methods are used. The weighted methods are usually more effective but prone to violations of the InSIDE assumption. 6.) The penalized weighted median method calculates median based estimators under penalization of weights of genetic variants with heterogenous effect estimates and is especially valuable in scenarios with high heterogeneity [4]. 7.) The MR-PRESSO method performs heterogeneity tests for each variant to detect outliers, which are consequently deleted for the calculation of outlier-corrected IVW estimates [16]. The raw estimate is similar to the IVW method. 8.) Similarly, MR-Lasso introduces an intercept for each variant representing the direct pleiotropic effect of each variant on the outcome. Intercepts are subject to lasso penalization to approximate intercepts of valid terms towards zero. An IVW estimate is calculated for valid instruments only (with an intercept of zero) [12]. Both methods, MR-PRESSO and MR-Lasso work efficient with few invalid instruments but may be less valuable with a large number of pleiotropic variants. 9.) The contamination mixture method builds a mixture model based on the categorization of instruments as valid and invalid and provides highly efficient (meaning high power) calculation of estimates even in the presence of invalid instrumental variables and high heterogeneity [5]. 10.) The MR-RAPS method models pleiotropic terms as random effects normally distributed around zero and downweighs outliers [19]. This method is especially valuable in the case of balanced pleiotropy.

**Methods 2 - Software**

Univariable MR studies were conducted with the TwoSampleMR package (version 0.5.7, [8]), the MendelianRandomization package (version 0.7.0, [18]) for the MR-Lasso and contamination mixture method as well as with the MRPRESSO (version 1.0, [16]) and the mr.raps (version 0.4.1, [19]) R-packages. For MVMR analyses, the MVMR extension of the MendelianRandomization package as well as the MVMR (version 0.4, [14]) package were used. Graphics were built with R packages ggplot2 (version 3.4.2) and forestploter (version 1.1.0). All analyses were conducted using R version 4.3.0 on RStudio (version 2023.03.0+386). Power analyses considering the sample size, the ratio of cases of the outcome GWAS, and the proportion of variance explained by the instrumental variable for each exposure were performed using the online mRnd tool (<https://shiny.cnsgenomics.com/mRnd/> see [3]). The proportion of the variance explained of the instrumental variable used for each exposure were calculated according to the formula by Shim, et al. [15].

**Methods 3 – Measurement/calculation of bioavailable testosterone**

In the exposure GWAS used in our MR study [13], total testosterone was measured by one step competitive analysis on a Beckman Coulter Unicel Dxl 800. Bioavailable testosterone was calculated via the Vermeulen equation accounting for SHBG (by two step sandwich immunoassay analysis on a Beckman Coulter Unicel Dxl 800) and albumin (measured on a Beckman Coulter AU5800). For sex-specific analyses, participants with testosterone levels below the lower detectable limit were excluded, resulting in 177,782 male and 188,507 female participants. For sex-combined genetic instruments, testosterone levels below the lower detectable limit were set to 0.3.

**Methods 4 – Exposure variables and populations included in MVMR studies**

MVMR analyses were conducted including BMI, birth weight, and SHBG as exposure variables. BMI was included as a result of the PhenoScanner search which revealed that several SNPs in the univariable MR analysis were either directly associated with weight and body composition or indirectly related with metabolic changes which itself are closely related to BMI. In addition, the pre-existing literature already has shown strong associations of BMI and other weight-related measures with testosterone as well as ADHD, making BMI a likely source of pleiotropy in our study. For BMI, summary statistics of a GWAS on 806,834 individuals (434,794 females) of European ancestry was used [11]. Birth weight was included as genetic variants of the IV were either directly associated with birth weight or associated, e.g. with arterial hypertension, itself strongly associated with low birth weight [1]. Additionally, birth weight has consistently been associated with ADHD in epidemiological studies [7], making it a likely source for pleiotropy. For this exposure, a European-only birth weight GWAS of up to 298,142 participants provided by Warrington, et al. [17] was used. SHBG was selected as a potential covariate as it shares a large genetic overlap with (bioavailable) testosterone [9, 13].

*BMI*

To include BMI as an exposure of interest, the GWAS by Pulit, et al. [11] was used. The GWAS for BMI was based on 806,834 individuals (434,794 females) of European ancestry. Data was either derived from the UK Biobank or generated by the Genetic Investigation of ANthropometric Traits (GIANT) consortium. For GWAS, only the SNP array used for genotype sampling was used as a covariate.

*Birth Weight*

For birth weight, summary statistics of the European-only birth weight GWAS of up to 298,142 participants provided by Warrington, et al. [17] was used. This GWAS is the result of a meta-analysis on 217,397 white European participants from the UK Biobank and 80,745 individuals from the Early Growth Genetics (EGG) Consortium which comprises 35 studies from the United States, Australia, and Europe. For UK Biobank participants, data collection based on self-reports of own birth weight of adults. Individuals with a birth weight <2.500g or > 4.500g or multiple births were excluded. For participants of the studies of the EGG consortium, birth weights were either derived from medical records or self-reports, usually individuals with pre-term delivery or multiple births were excluded. In both studies, birth weights were z-transformed for males and females to account for sex-specific effects. The genotyping array and year of birth as well as the gestational age (only partially available in studies of the EGG consortium) were included as covariates of the resulting GWAS.

### Supplementary Results

**Results 1 - MR study on bioavailable testosterone on ADHD (sexes-combined)**

Details on effect estimates as well as on tests of heterogeneity and MR-PRESSO global test for pleiotropy can be found in the main document. MR-PRESSO identified three pleiotropic SNPs (rs1811450, rs28929474 and rs4431046) which were excluded from further analysis, resulting in 107 SNPs defining the IV. The SNP rs1811450 is located in the intron region of the METTL15 gene which regulates the methylation of mitochondrial 12S rRNA and thus has widespread phenotype associations e.g. with body size or whole body fat-free mass. rs28929474 is a missense mutation in the SERINA1-Gen which encodes for a serine protease inhibitor with defects leading to Alpha-1 antitrypsin deficiency, resulting in chronic liver failure and lung emphysema. rs4431046 is in the intron region of the PPP2R3A gene that plays a role in the control of cell growth and division. Phenotype associations of this SNP include BMI and coronary heart disease.

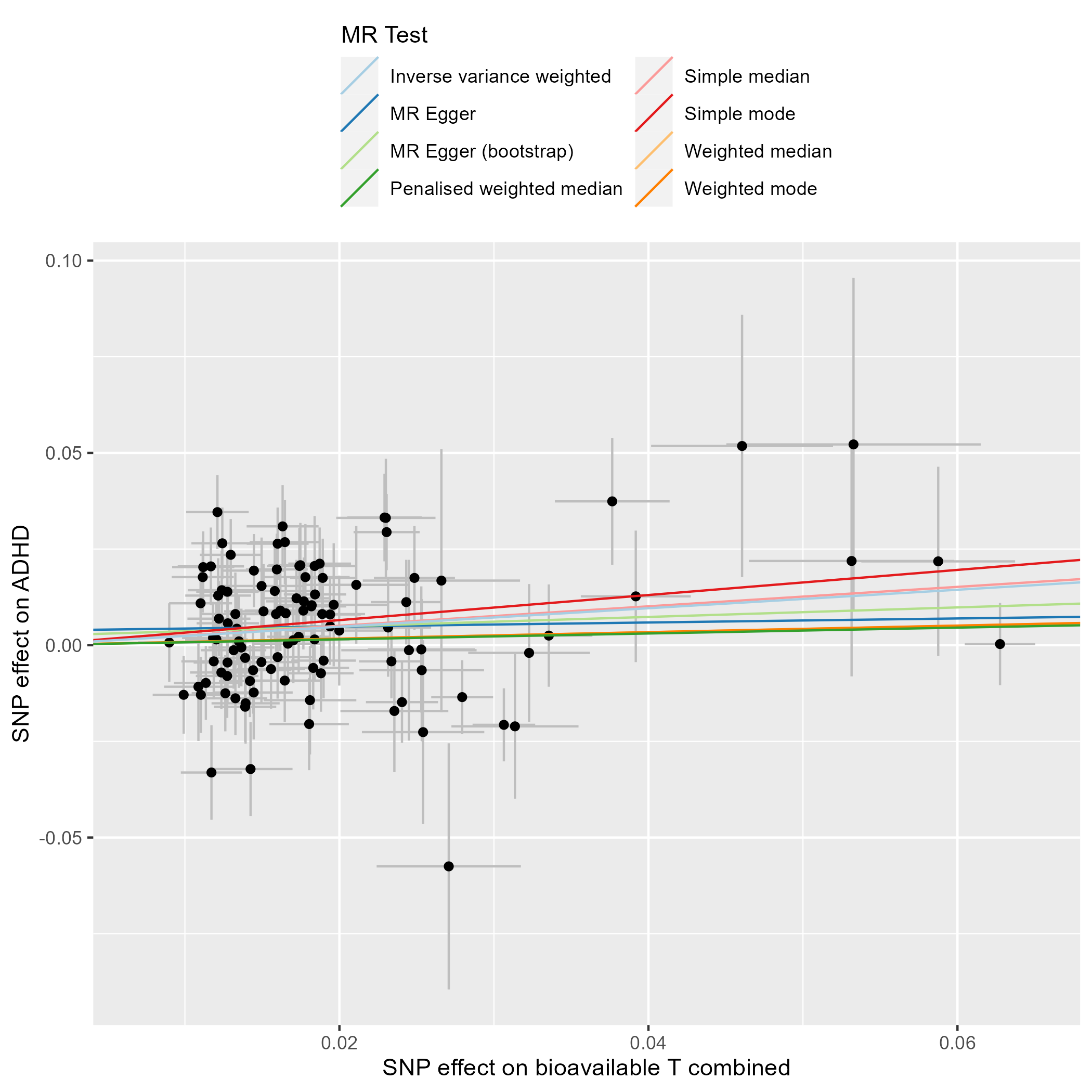

**Supplementary Figure S1. Scatter plot of the SNPs utilized in the univariable MR analysis (sexes combined).** This scatter plot illustrates the beta effect estimates of each genetic variant included in the analysis on the exposure variable (bioavailable testosterone, [13]) and the outcome variable (ADHD, [6]).

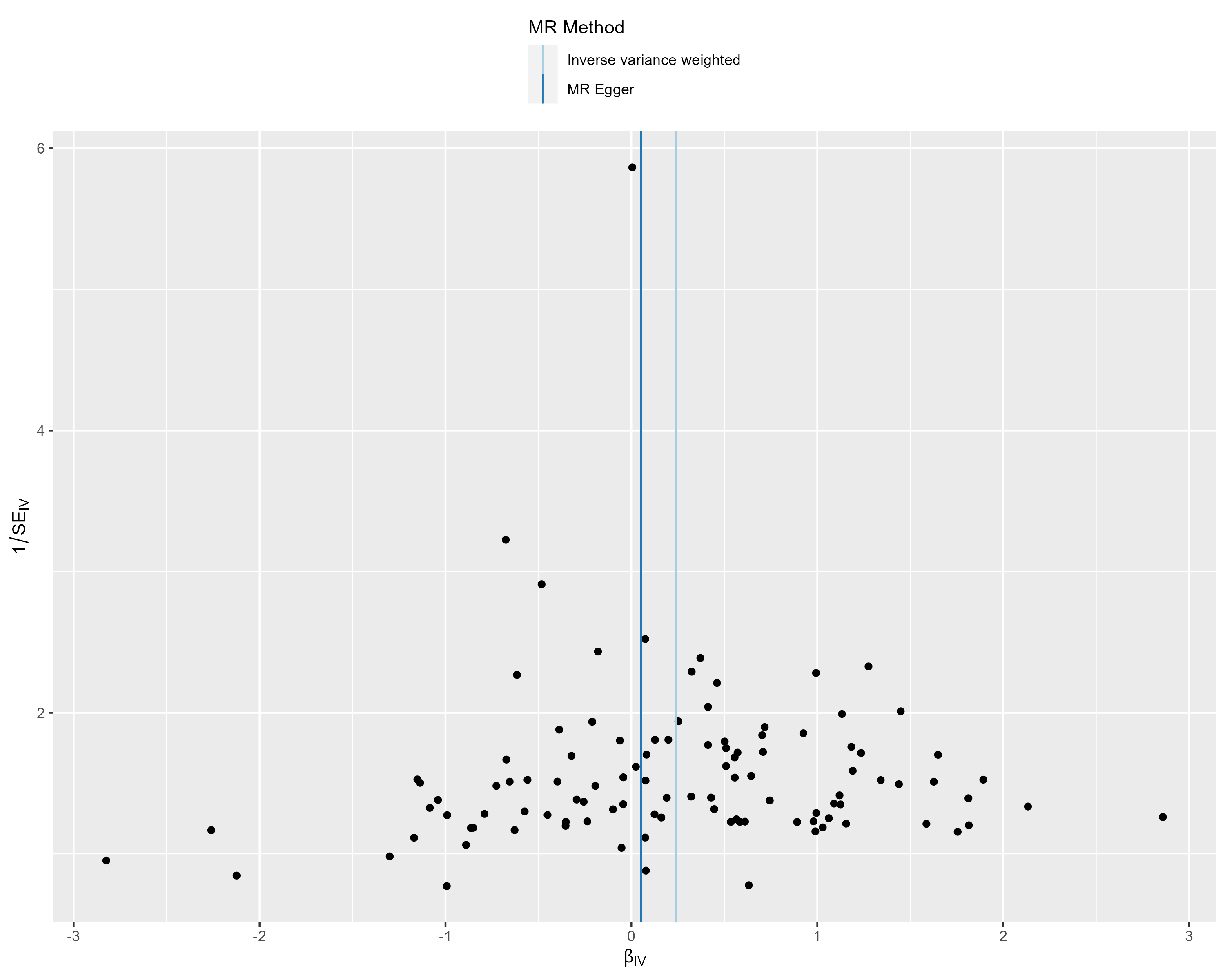

**Supplementary Figure S2. Funnel plot of the SNPs utilized in the univariable MR analysis (sexes combined).** This funnel plot illustrates the relationship between the effect estimate (β_IV_) of each genetic instrument and inverse of the standard error (SE_IV_) as a measure of their precision. Strong instruments (i.e. instruments with a high 1/ SE_IV_ ratio) are approximately scattered around the IVW and MR Egger estimates, indicating no major source for directional pleiotropy.

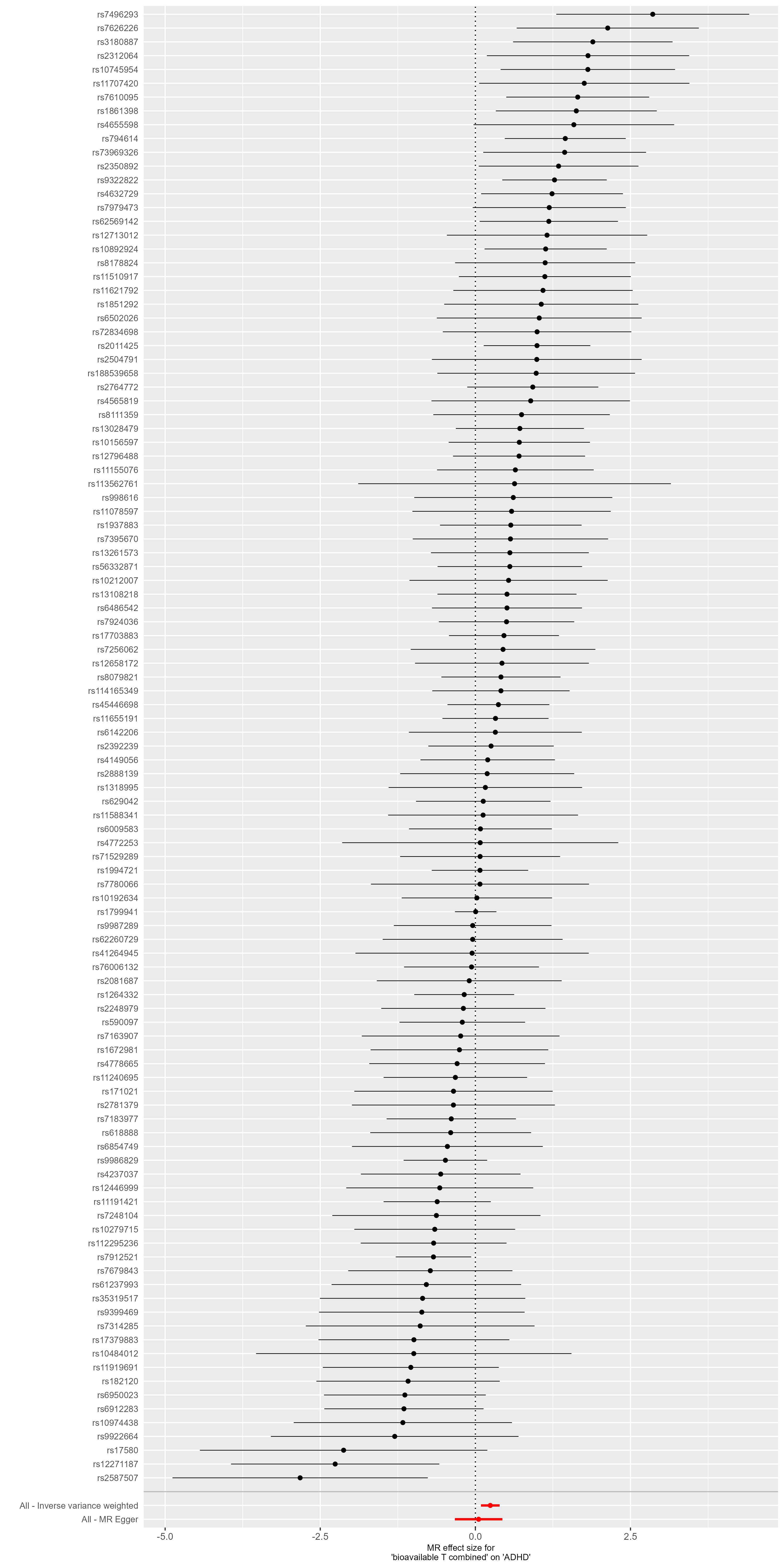

**Supplementary Figure S3. Forest plot of the SNPs utilized in the univariable MR analysis (sexes combined).** This forest plot illustrates the MR effect estimates for each SNP on ADHD in single SNP MR analyses. No major outliers were found.

**Results 2 - MR study on bioavailable testosterone on ADHD (males only)**

Details on effect estimates as well as on tests of heterogeneity and the MR-PRESSO global test for pleiotropy can be found in the main document.

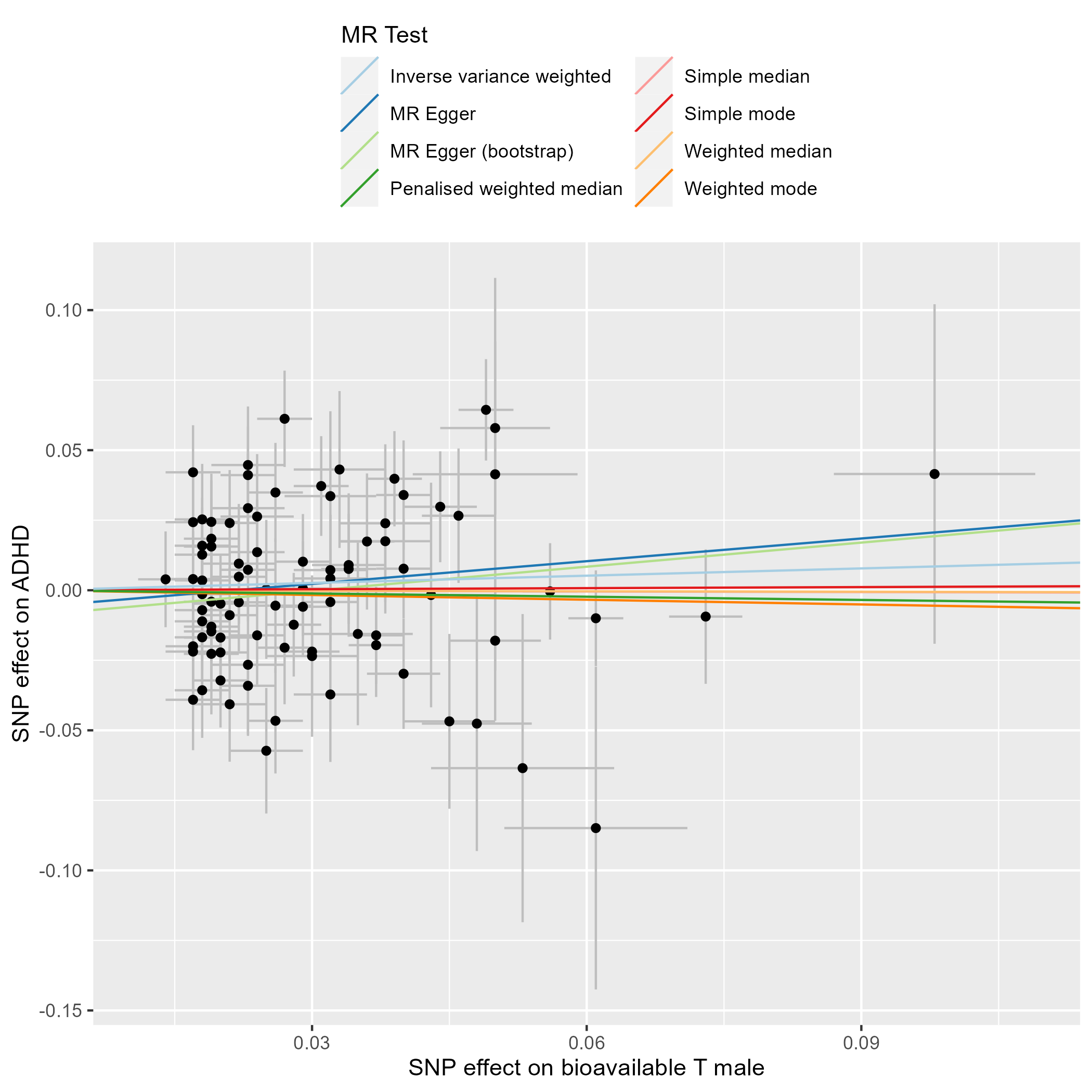

**Supplementary Figure S4. Scatter plot of the SNPs utilized in the univariable MR analysis (males only).** This scatter plot illustrates the beta effect estimates of each genetic variant included in the analysis on the exposure variable (bioavailable testosterone in males, [13]) and the outcome variable (ADHD in males, [10]).

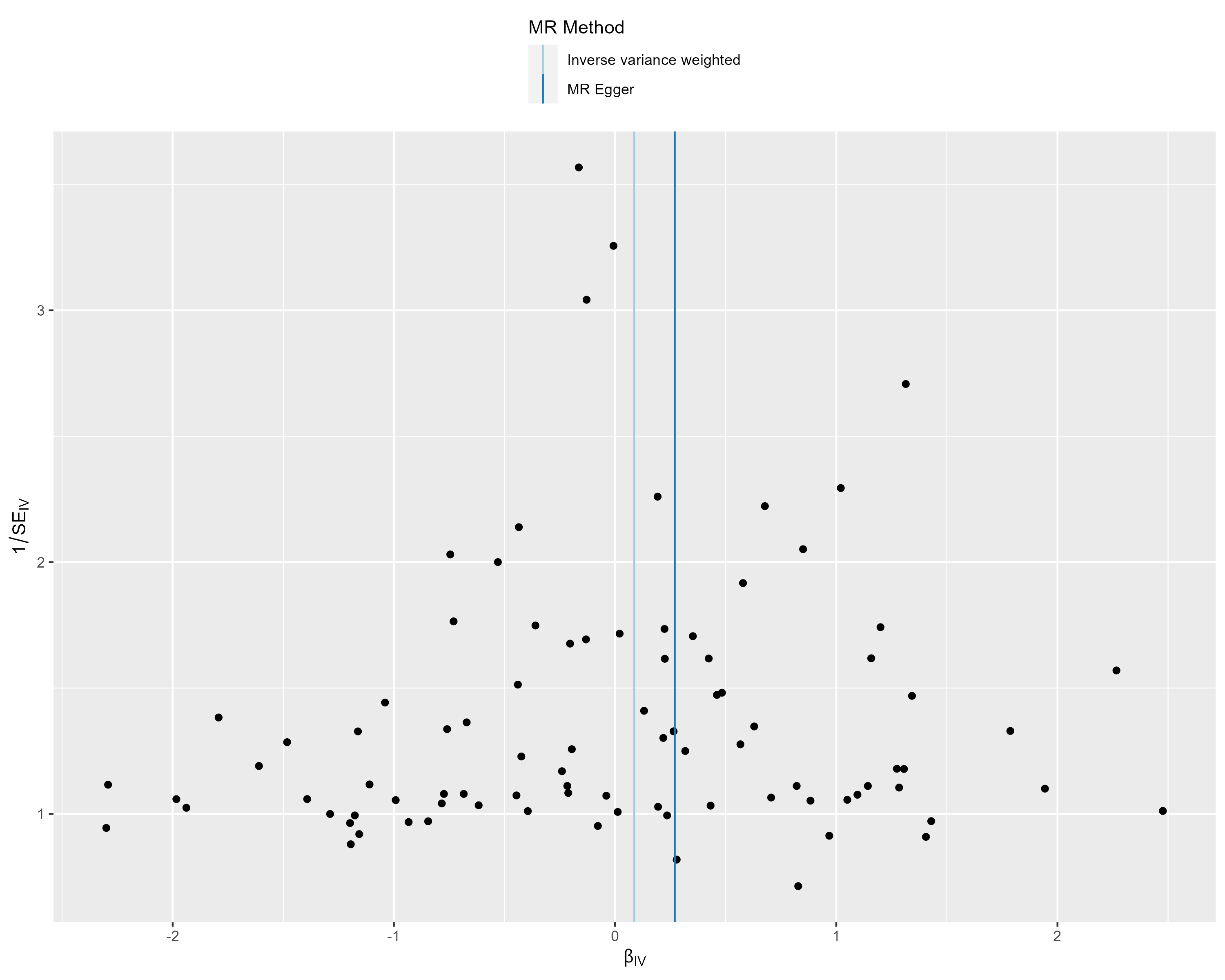

**Supplementary Figure S5. Funnel plot of the SNPs utilized in the univariable MR analysis (males only).** This funnel plot illustrates the relationship between the effect estimate (βIV) of each genetic instrument and inverse of the standard error (SEIV) as a measure of their precision.

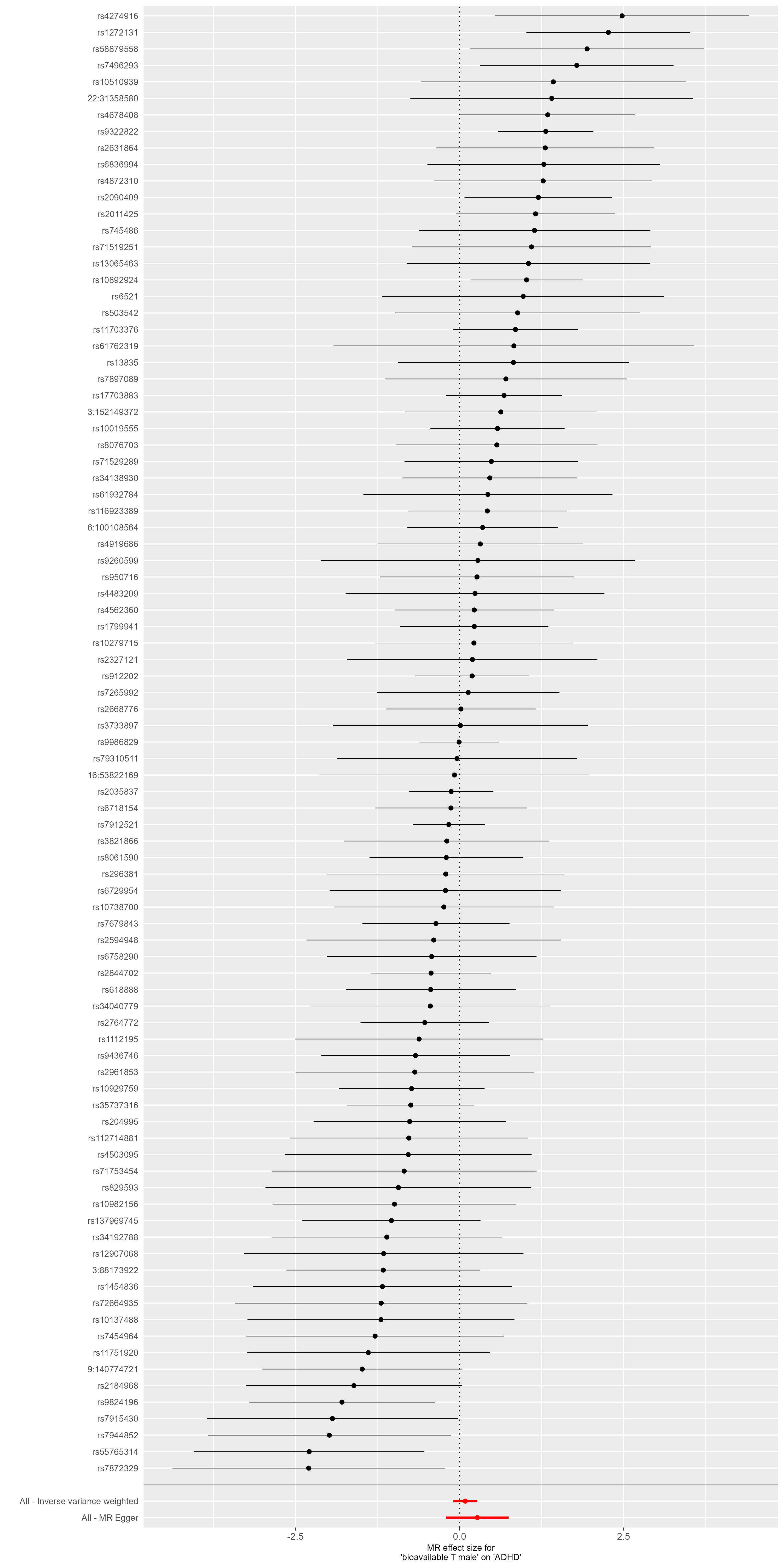

**Supplementary Figure S6. Forest plot of the SNPs utilized in the univariable MR analysis (males only).** This forest plot illustrates the MR effect estimates for each SNP on ADHD in single SNP MR analyses.

**Results 3 - MR study on bioavailable testosterone on ADHD (females only)**

Details on effect estimates as well as on tests of heterogeneity and MR-PRESSO global test for pleiotropy can be found in the main document.

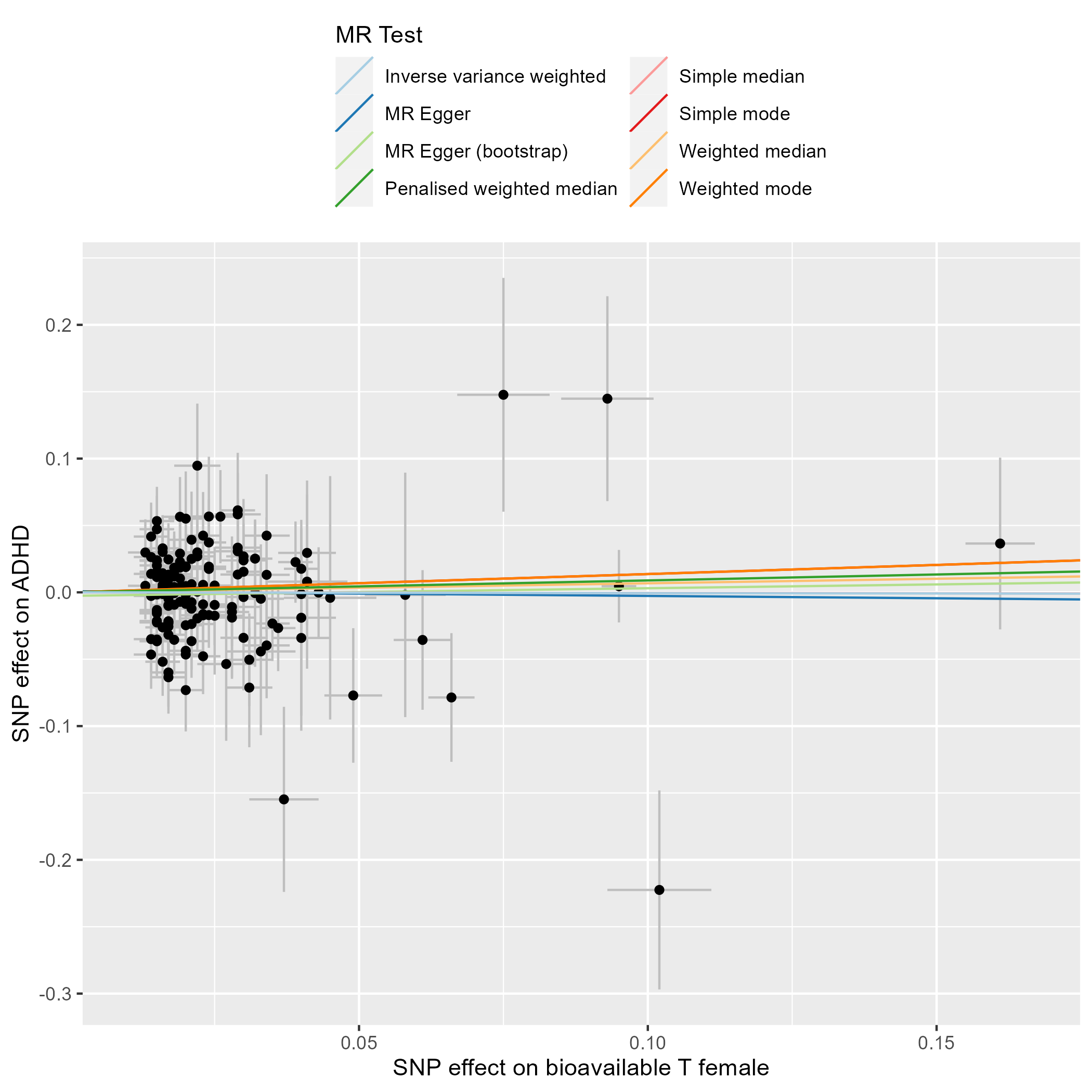

**Supplementary Figure S7. Scatter plot of the SNPs utilized in the univariable MR analysis (females only).** This scatter plot illustrates the beta effect estimates of each genetic variant included in the analysis on the exposure variable (bioavailable testosterone in females, [13]) and the outcome variable (ADHD in females, [10]).

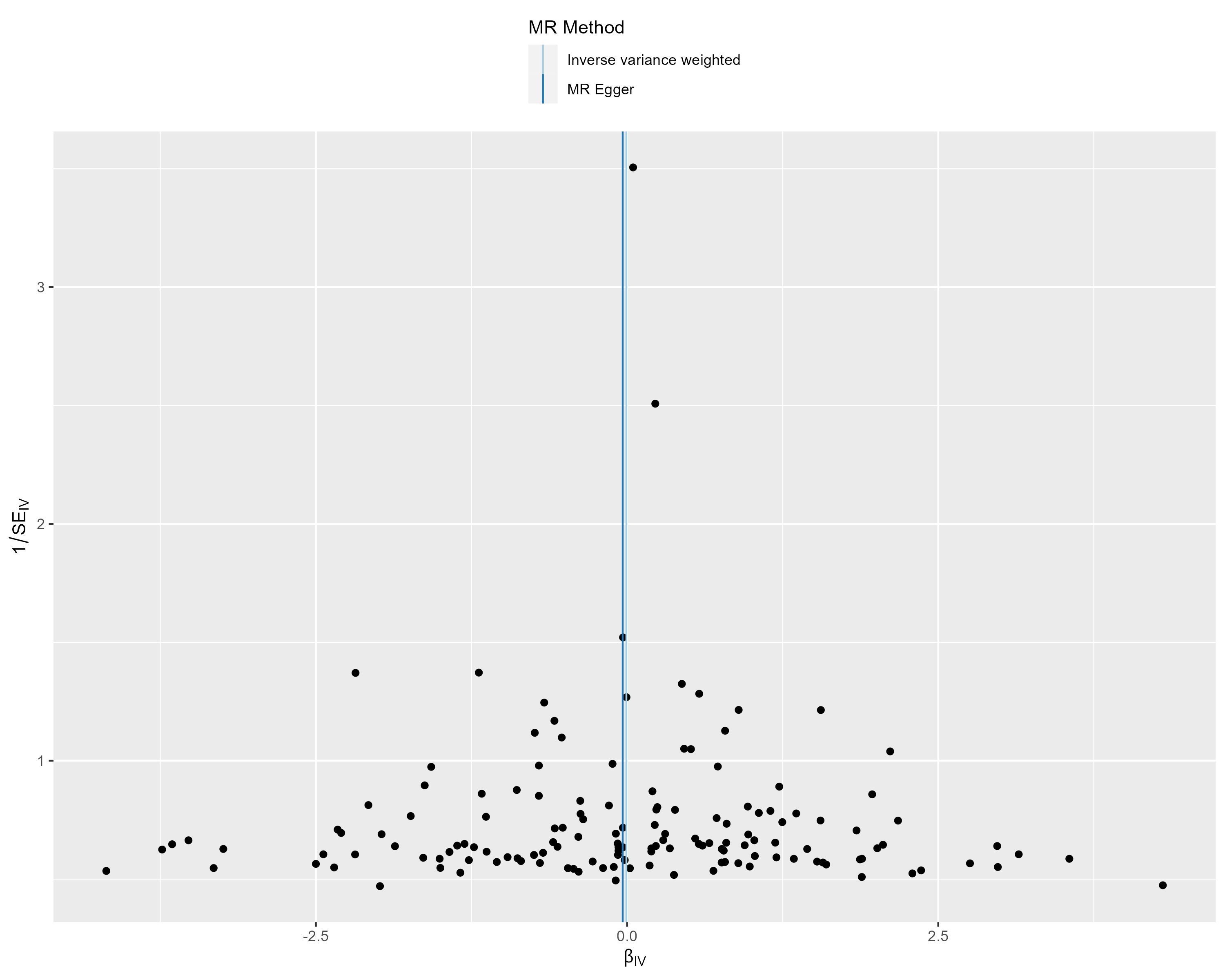

**Supplementary Figure S8. Funnel plot of the SNPs utilized in the univariable MR analysis (females only).** This funnel plot illustrates the relationship between the effect estimate (βIV) of each genetic instrument and inverse of the standard error (SEIV) as a measure of their precision.

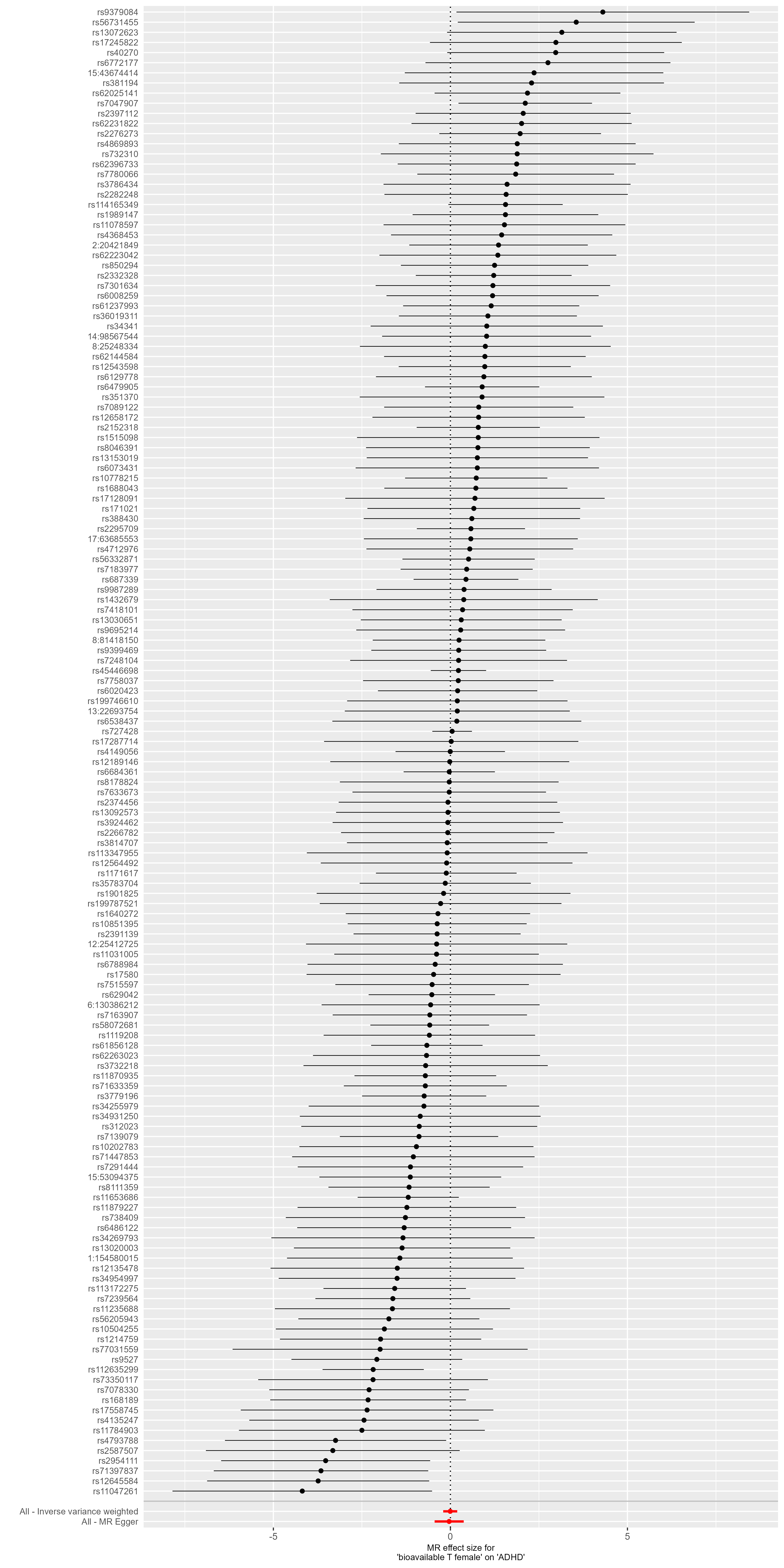

**Supplementary Figure S9. Forest plot of the SNPs utilized in the univariable MR analysis (females only).** This forest plot illustrates the MR effect estimates for each SNP on ADHD in single SNP MR analyses.

**Results 4 - Sensitivity analyses (sexes-combined analyses)**

To test whether the r^2^ threshold for proxy SNP search, the harmonization process or the exclusion of SNPs by MR-PRESSO biased the results of our study, two additional analyses were conducted i.) a univariable MR analysis of bioavailable testosterone on ADHD (sex-combined) without exclusion of pleiotropic SNPs as identified by MR-PRESSO including proxies only with r²>0.8 and otherwise unchanged methodology and ii.) a univariable MR analysis of bioavailable testosterone on ADHD (sex-combined) without exclusion of pleiotropic SNPs as identified by MR-PRESSO but with exclusion of ambiguous palindromic SNPs (meaning genetic variants with a minor allele frequency of ≥ 0.42 and thus potentially misalignment with harmonization) and otherwise unchanged methodology.

*Proxy selection limited to proxies with r²>0.8*

A more conservative cut-off of r²>0.8 lead to the identification of 15 proxies, leading to a total of 102 genetic variants included in the MR-analysis. This analysis lead to a significant positive finding of the contamination mixture method (estimated effec =0.67, lower-95%-CI=0.04, upper-95%-CI=0.93, p=0.031). All other analyses yielded similar results compared to the main study: while the IVW, MR-RAPS, median, MR-PRESSO and MR-Lasso methods revealed significantly positive effect estimates (range from 0.21 to 0.32, p values range from 0.001 to 0.009), the effect estimates based on the MR-Egger, weighted median, penalized weighted median and mode-based methods were not significant (estimates range from -0.14 to 0.36, p values between 0.210 and 0.457), with a significant Egger’s intercept (intercept 0.0077±0.007, p=0.041). The Q-statistic revealed heterogeneity (IVW: Q-statistic: 193.23, p=9.35x10^-8^; MR-Egger: Q-statistic: 185.27, p=4.72x10^-7^). The MR-PRESSO global test for pleiotropy was significant (197.78, p<3.3x10^-4^). Supplementary Figure 10 illustrates the effect estimates of this approach.

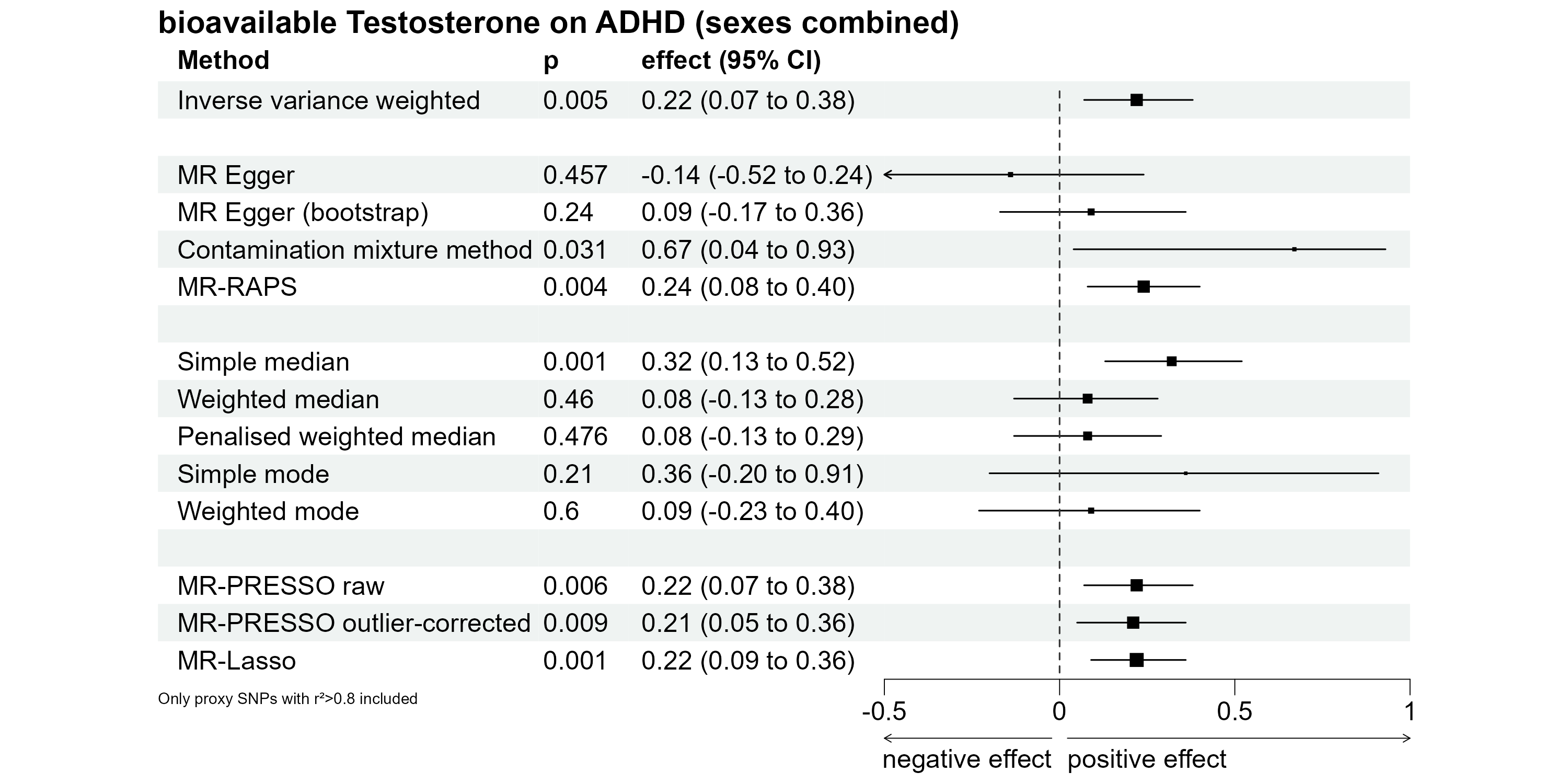

**Supplementary Figure S10. Effect estimates in sexes-combined MR analysis with proxy selection based on r²>0.8**

*Harmonization with exclusion of ambiguous palindromic SNPs*

For harmonization in primary analysis, it was assumed that all alleles are coded on the forward strand for palindromic SNPs. A more conservative approach in the harmonization process lead to the exclusion of four SNPs with intermediate minor allele frequencies, resulting in an instrumental variable consisting of 106 genetic variants. Using this approach, the simple median-based effect estimate was not significant (estimated effect=0.18, lower-95%-CI=-0.02, upper-95%-CI=0.38, p=0.180). The remaining results were similar to the results of the primary analysis, with significant positive effect estimates with IVW, MR-RAPS, MR-PRESSO and MR-Lasso (range from 0.18 to 0.21, p values range from 0.002 to 0.022), while MR-Egger, all median- and mode-based effect estimates as well as the contamination mixture method revealed non-significant findings (effect estimates range from 0.07 to 0.28, p values range from 0.082 to 0.638). Q-statistic revealed a significant amount of heterogeneity (IVW: Q-statistic: 235.27, p=5.52x10^-12^; MR-Egger: Q-statistic: 234.78, p=4.18x10^-12^). Egger’s intercept did not show evidence for significant directional pleiotropy (intercept = 0.0019±0.0040, p=0.641). Supplementary Figure 11 illustrates the effect estimates of this approach.

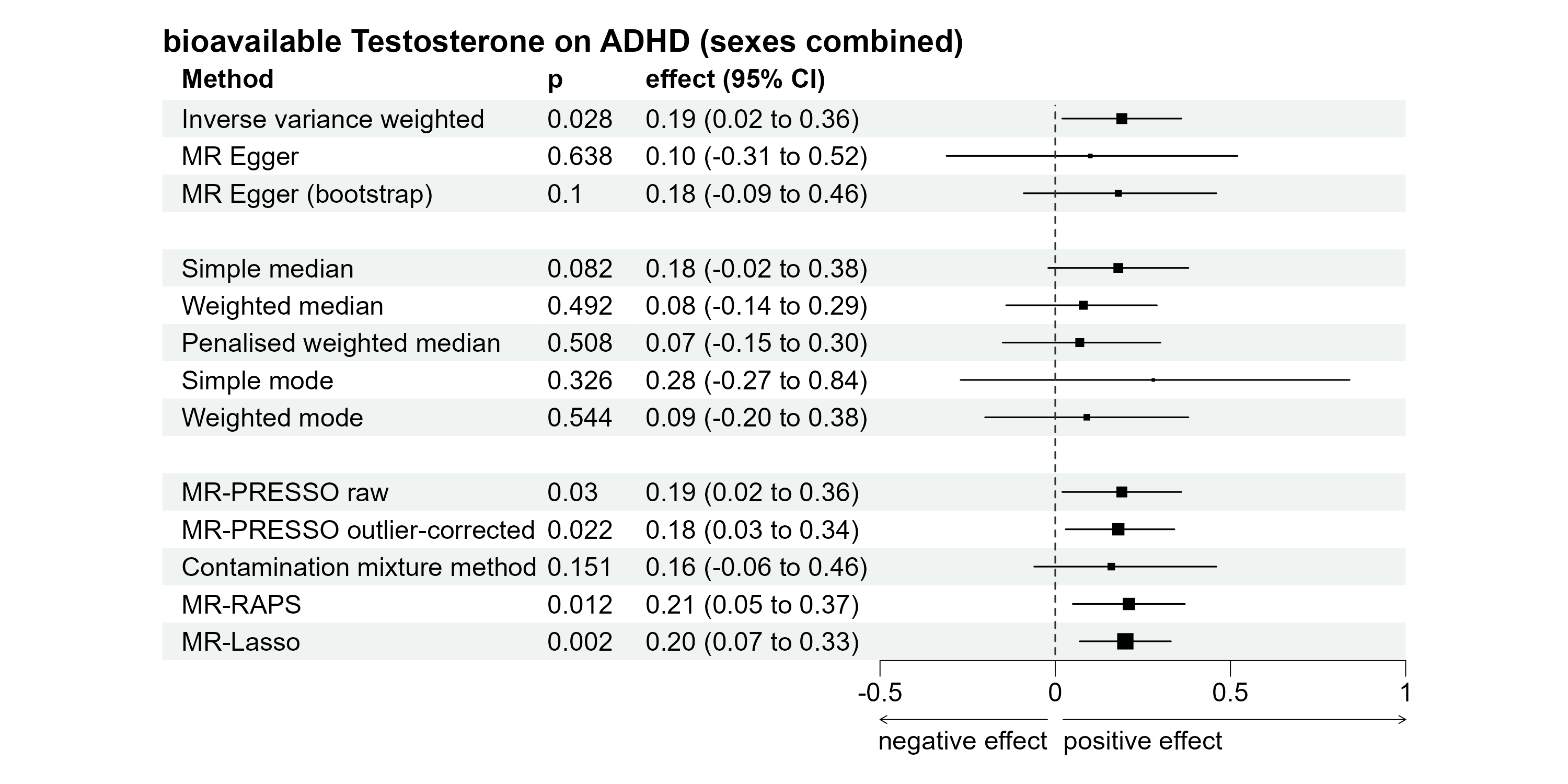

**Supplementary Figure S11. Effect estimates in sexes-combined MR analysis with exclusion of palindromic SNPs with intermediate allele frequencies.**

**Results 5 - Sensitivity analyses (sex-specific analyses)**

The effect-allele frequency was not given in the sex-specific GWAS on ADHD. Therefore, the effect-allele frequency could not be utilized for harmonization of palindromic SNPs in sex-specific analyses. We recalculated the sex-specific MR-analyses with instrumental variables excluding palindromic SNPs, leading to an instrumental variable consisting of 74 SNPs in males and of 129 SNPs in females (both with proxies). In the male-specific MR-analysis, MR-Egger with bootstrapping now leads to a non-significant finding (b=0.24, p=0.095). All other analyses provided similar effect estimates as the primary analyses (Supplemental Figures S12 and S13).

*Exclusion of palindromic SNPs in males*

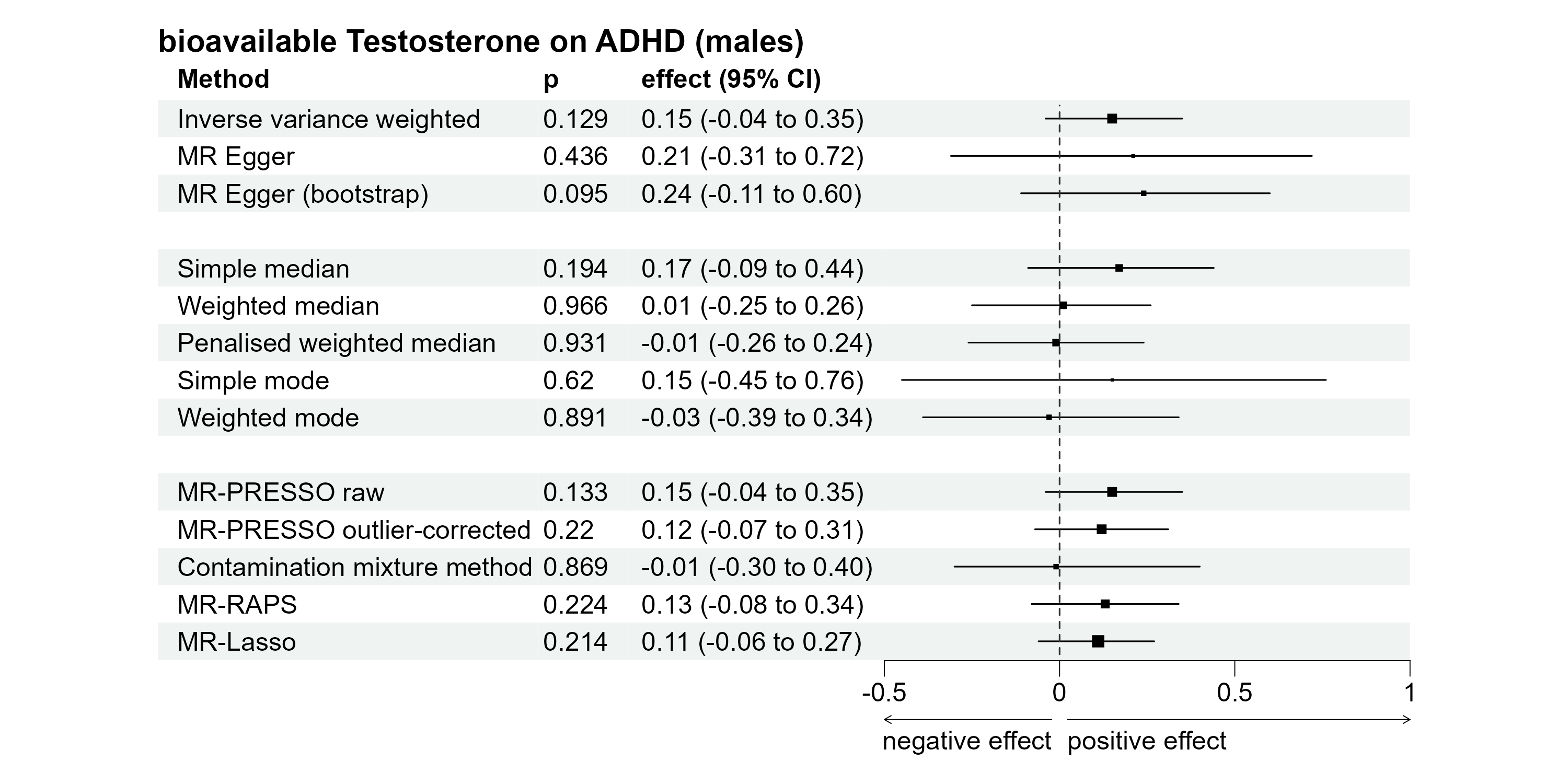

**Supplementary Figure S12. Effect estimates in males-only MR analysis with exclusion of palindromic SNPs.**

*Exclusion of palindromic SNPs in females*

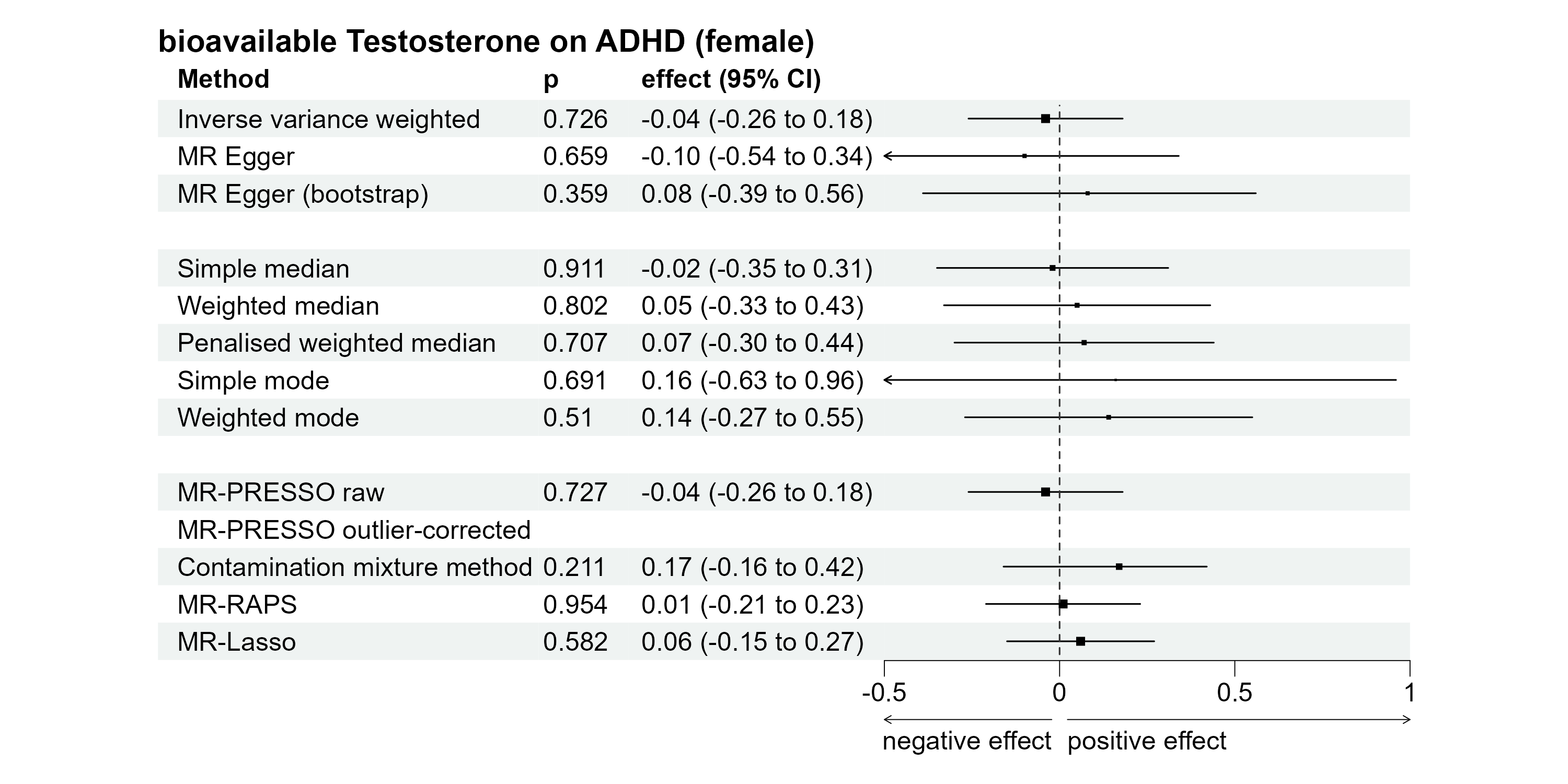

**Supplementary Figure S13. Effect estimates in females-only MR analysis with exclusion of palindromic SNPs.**

**Results 6 - Sensitivity analyses (MVMR)**

Details on effect estimates as well as on tests of heterogeneity and MR-PRESSO global test for pleiotropy can be found in the main document. MR-PRESSO identified two pleiotropic SNPs (rs1811450 and rs7496293) which were excluded in consecutive analyses. For the same reasons, rs1811450 was already excluded in primary analyses, the description of the pleiotropic associations of rs1811450 can be found in Supplementary Results 1. rs7496293 is located in the intron region of the gene encoding for the transcription factor 12 which is expressed in many tissues. Phenotype associations include BMI and platelet count. To assess whether the exclusion of these SNPs biased our results, the analyses were recalculated without exclusion of pleiotropic SNPs. The results, which replicated the findings of the primary analysis, can be found in Supplemental Figure S14. For the second MVMR model including bioavailable testosterone, SHBG (BMI adjusted), and birth weight as exposure variables, MR-PRESSO identified two pleiotropic SNPs (rs1811450, rs4431046) with pleiotropic associations. rs1811450 and rs4431046 were already excluded in primary analysis, further details on both SNPs can be found in Supplementary Results 1.

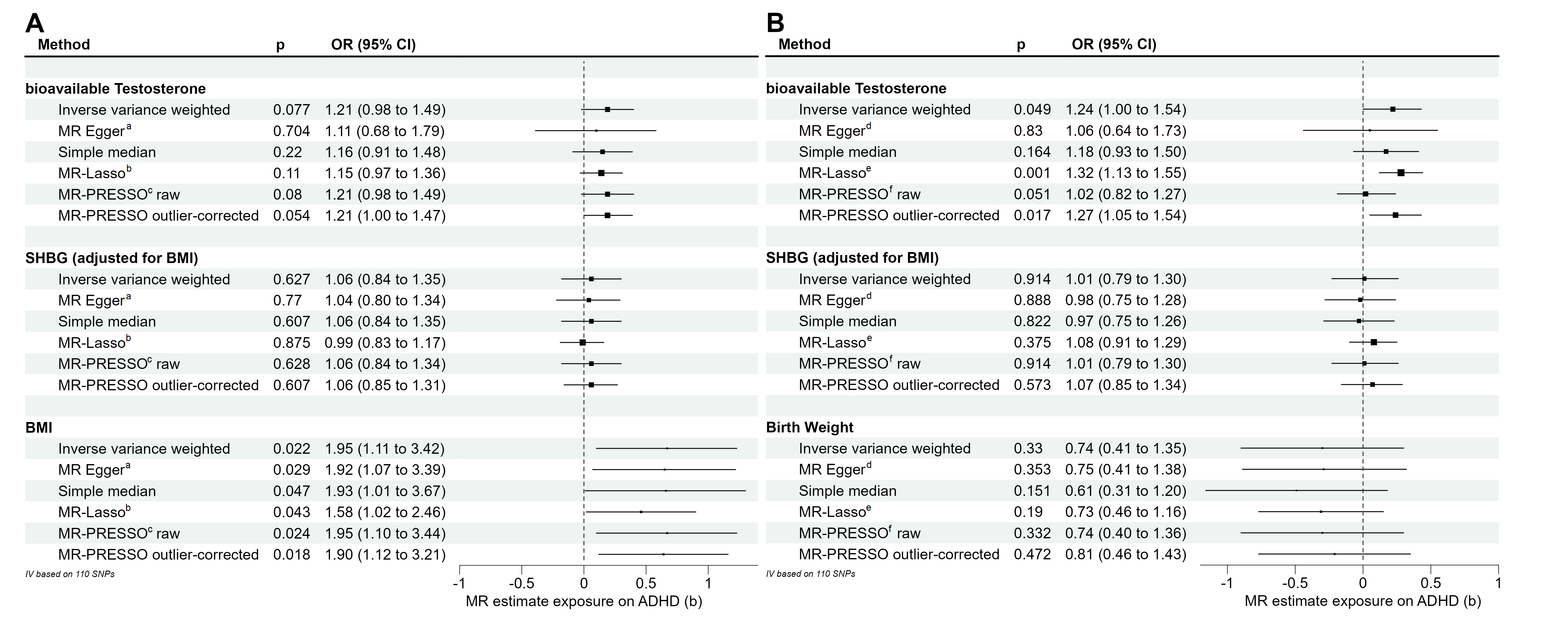
 **Supplementary Figure S14 – MVMR analysis without exclusion of pleiotropic SNPs.** This Figure depicts the effect estimates of the two MVMR analyses conducted without exclusion of pleiotropic SNPs as identified by the MR-PRESSO method. Figure S14A depicts the results of the MVMR analysis with bioavailable testosterone, SHBG (BMI-adjusted) and BMI. Figure S14B depicts MVMR analysis with bioavailable testosterone, SHBG (BMI-adjusted) and birth weight. ^a^ Eggers-intercept did not show evidence for significant directional pleiotropy (intercept=0.002, se=0.004, p=0.675). ^b^ The MR-Lasso method identified 87 valid instruments (tuning parameter=0.171). ^c^ MR-PRESSO global test for pleiotropy was significant (246.7, p < 4x10^-4^). ^d^ Eggers-intercept=0.003, se=0.004, p=0.482). ^e^ The MR-Lasso method identified 90 valid instruments (tuning parameter=0.182). ^c^ The MR-PRESSO global test for pleiotropy was significant (257.1, p < 3x10^-4^).
